# Estimation and optimal control of the multi-scale dynamics of the Covid-19

**DOI:** 10.1101/2021.03.04.21252880

**Authors:** David Jaurès Fotsa-Mbogne, Stéphane Yanick Tchoumi, Yannick Kouakep-Tchaptchie, Vivient Corneille Kamla, Jean-Claude Kamgang, Duplex Elvis Houpa-Danga, Samuel Bowong-Tsakou, David Bekolle

## Abstract

This work aims at a better understanding and the optimal control of the spread of the new severe acute respiratory corona virus 2 (SARS-CoV-2). We first propose a multi-scale model giving insights on the virus population dynamics, the transmission process and the infection mechanism. We consider 10 compartments in the human population in order to take into accounts the effects of different specific mitigation policies: susceptible, infected, infectious, quarantined, hospitalized, treated, recovered, non-infectious dead, infectious dead, buried. The population of viruses is also partitioned into 10 compartments corresponding respectively to each of the first nine human population compartments and the free viruses available in the environment. Indeed, we have human to human virus transmission, human to environment virus transmission, environment to human virus transmission and self infection by susceptible individuals. We show the global stability of the disease free equilibrium if a given threshold *𝒯*_0_ is less or equal to 1 and we provide how to compute the basic reproduction number *ℛ*_0_. A convergence index *𝒯*_1_ is also defined in order to estimate the speed at which the disease extincts and an upper bound to the time of extinction is given. The existence of the endemic equilibrium is conditional and its description is provided. We evaluate the sensitivity of *ℛ*_0_, *𝒯*_0_ and *𝒯*_1_ to control parameters such as the maximal human density allowed per unit of surface, the rate of disinfection both for people and environment, the mobility probability, the wearing mask probability or efficiency, and the human to human contact rate which results from the previous one. Except the maximal human density allowed per unit of surface, all those parameters have significant effects on the qualitative dynamics of the disease. The most significant is the probability of wearing mask followed by the probability of mobility and the disinfection rate. According to a functional cost taking into consideration economic impacts of SARS-CoV-2, we determine and discuss optimal fighting strategies. The study is applied to real available data from Cameroon and an estimation of model parameters is done. After several simulations, social distancing and the disinfection frequency appear as the main elements of the optimal control strategy.

## 1 Introduction

According to the World Health Organization^1^ (WHO), the severe acute respiratory syndrome corona virus 2 (SARS-CoV-2) previously known as new corona virus 2019 (2019-nCoV) is responsible of an infectious disease called Covid-19 [49]. In 1937, corona viruses were first identified as infectious bronchitis viruses with which birds suffered that could devastate poultry stocks. Today, the viruses are the cause of the common cold in 15% to 30% of all cases. In the past 70 years, researchers have found camels, cattle, cats, dogs, horses, mice, pigs, rats and turkeys that were infected with corona viruses^2^. Older people, and those with underlying medical problems like cardiovascular diseases, diabetes, chronic respiratory diseases, and cancer are more likely to develop serious illness. The Centers for Disease Control and Prevention ^3^ (CDC) affirms that the most common ways the virus spreads from an infected person to healthy people around them is when they cough or sneeze and release viral particles into the air and through touching, hands shaking and others forms of close personal contact. When healthy people touch objects or surfaces on which there are viral particles, then touch their eyes, nose or mouth before washing their hands, the virus can spread. In some rare cases fecal contamination can cause the virus to spread as well. The best way to prevent and slow down transmission is being well informed about the SARS-CoV-2, the disease, its causes and how it spreads. Each person protects himself and others from infection by hands washing or using an alcohol based rub frequently and not touching his face. The SARS-CoV-2 spreads primarily through droplets of saliva or discharge from the nose when an infected person coughs or sneezes, so it is important to also practice respiratory etiquette (for example, by coughing into a flexed elbow). Notice that at this time, there are no specific vaccines or treatments for Covid-19. Depending on its location the SARS-CoV-2 can leave up to 7 days out of human body [10]. Modeling, analyzing models, estimating risks and forecasting the potential spread of the disease in population appears very useful for decision makers [32].

Recently, numerous papers (published or not) appeared in order to contribute to the fight against the pandemic Covid-19. Those papers can be organized in three groups. The first group addressed the problem of forecasting the disease in order to help decision makers to better evaluate the logistic challenges they will face [1, 3, 4, 16, 15, 24, 25, 28, 29, 37, 38, 39, 41, 45, 47, 48, 51, 61, 62]. The second category of papers focused on evaluating the effectiveness of mitigations measures prescribed by the WHO and different governments in order to define better fighting strategies [5, 9, 12, 13, 14, 29, 34, 37, 41, 44, 45, 46, 50, 53, 55, 54, 60, 62]. The last set of papers studied the social and the economical impacts of the pandemic [35, 59]. As we can observe in the literature, the wide part of models only consider person to person disease transmission. Of course it is important, but indirect transmission by environment may also be considered. On the other hand the virus population dynamics appears also important and should be explicitly studied. Indeed, when an individual gets in contact with the virus he is not directly infected. He really becomes exposed when the virus penetrates his organism by oral or respiratory ways. Hence, the exposition is mainly indirect in terms of human to human or human to environment contacts. This consideration justifies the regular disinfection of individuals and the environment as recommended by public health agencies. Following the authors in [19, 20, 21, 22, 23, 43], it is important to build and understand models that link the within-host dynamics and population level dynamics of infectious. The prediction of the behavior of Covid-19 is very difficult because mitigation strategies are permanently changing. So it could be more efficient given an initial situation to propose optimal control strategies that will lead to disease extinction at a time to be determined. Hence, this work proposes efficient and low cost control strategies against Covid-19.

The paper is organized as follows. A model is described in subsection 2.1 and its asymptotic behavior is studied in subsection 2.2 depending on some critical thresholds we define. In subsection 3.1, we estimate the parameters of the model according to the real available data from Cameroon. A sensitivity analysis of critical thresholds depending on some control parameters is carried out in subsection 3.2. Section 4 is devoted to the design and the computation of optimal control strategies according to different constraints. Illustrative simulations are carried out in section 4.2. The algorithm which computes the optimal control is given in appendix C. The proofs of different theoretical results in appendix B. The paper ends with a conclusion in section 5.

## 2 The model and its general features

### 2.1 Description of the model

The disease dynamics can be viewed under two angles. Indeed, there are human dynamics on the first hand and the viruses dynamics on humans or in the environment on the other. We consider ten compartments in the human population per unit area (*m*^−2^): Susceptible (*S*), Infected (*E*), Infectious (*I*), Quarantined (*Q*), Hospitalized (*H*), Treated (*T*), Recovered (*R*), Non-Infectious dead (*D*), Infectious dead (*D*_*I*_), Buried (*B*). Each individual is assumed to have an external viral load on his body while there are free viruses in the environment. Again, we emphasis on the fact that getting in contact with viruses does not mean to be infected, but the infection occurs when the viruses penetrate the organism by oral or respiratory ways. Thus, we have ten compartments for virus population per individual status group located in a unitary surface or per unit area of environment (*virus × m*^−2^): global load in susceptible compartment (*V*_*S*_), global load in infected compartment (*V*_*E*_), global load in infectious compartment (*V*_*I*_), global load in quarantined compartment (*V*_*Q*_), global load in hospitalized compartment (*V*_*H*_), global load in treated compartment (*V*_*T*_), global load in recovered compartment (*V*_*R*_), global load in non-infectious dead compartment (*V*_*D*_), global load in infectious dead compartment 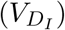, global free load in the environment (*V*_*F*_). The viral load is transfered from an individual being in compartment *i* to the environment (transfer of type I: *τ*_1,*i*_) or conversely from the environment to an individual being in compartment *i* (transfer of type II: *τ*_2,*i*_). Additionally, there are viral load transfers among individuals when they are in contact (transfer of type III: *τ*_3,*i*_). Notice that Infectious, quarantined, hospitalized and treated individuals produce new viruses at a rate *π*_*i*_ with *i* indexing a specific compartment. A susceptible individual becomes infected when a virus passes from the outside of his body to the inside (transfer of type IV: *τ*_4_). Infected persons die at a rate *µ* or become infectious with a rate *α*_1_. An infectious case is reported at a rate *q* and dies at a rate *µ* + *d*. The probability for a reported infectious person to be asymptomatic and then put in quarantine is *p*, otherwise he is admitted to hospital. With a rate *α*_2_ an infectious asymptomatic person is admitted to hospital after complications. Infectious, quarantined and hospitalized individuals die at a rate *µ* + *d* or become treated at the respective rates of *γ*_1_, *γ*_2_ and *γ*_3_. A treated individual is out of danger but is still infectious and can die at a rate *µ*. Treated persons recover completely and become immune at a rate *γ*_4_. A recovered person losses his immunity at a rate */!* or dies at a rate *µ*. With a rate *ρ*_1_, a non-infectious dead person is buried while an infectious dead individual is buried at a rate *ρ*_2_. Indeed, infectious dead persons are potentially very contagious and it is recommended to bury them rapidly in order to limit contacts.

WHO and the Government of Cameroon recommend several control measures that can be explicitly listed here as follows:

- Limiting movements through a control parameter (*m*),
- Limiting the area of movements through a lock-down ratio 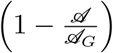,
- Limiting direct contacts with others through a control parameter (*τ*_3_),
- Frequently hands washing and disinfection through a control parameter (*ω*),
- Wearing mask and avoiding to touch the face through a control parameter (*u*).
- Social distancing which is implemented through limitation of group size per unit area to a maximum value (*κ*),

The disease is better contained if its spatial spreading is limited by lock-down. Multiplying movements increases the risk of being in contact with the virus from environment or from a person after a direct contact. By frequently hands washing and avoiding to touch the face (especially eyes, nose and mouth), the probability to get infected is clearly reduced. Wearing mask reduces both the risk of infection and the release of viruses by infectious, quarantined, hospitalized and treated individuals. We can summarize the dynamics of Covid-19 by the Figure 1.

**Figure 1.**
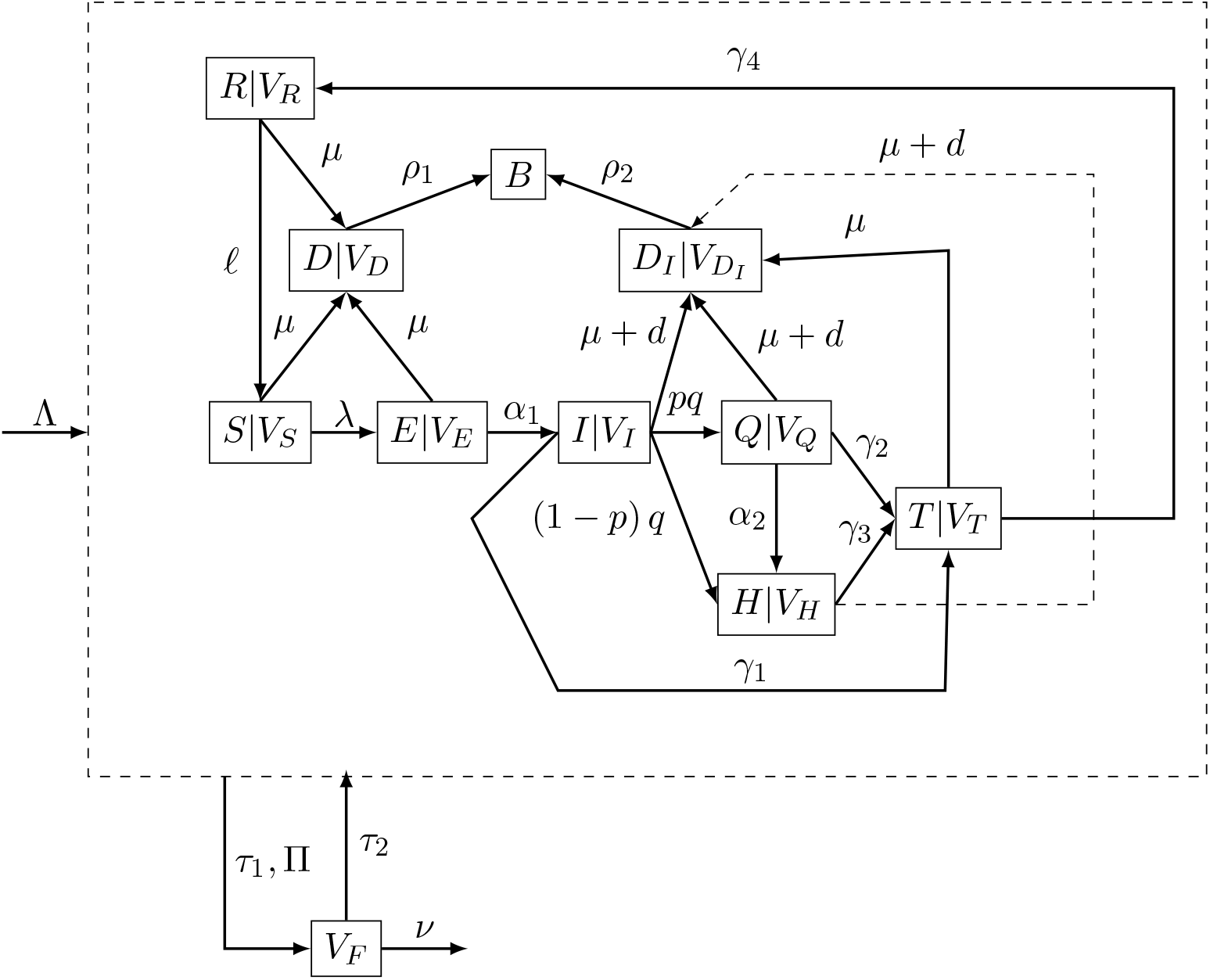
Flow chart of the Susceptible (*S*)-Exposed (*E*)-Infectious (*I*)-Quarantined (*Q*)-Hospitalized (*H*)-Treated (*T*)-Recovered (*R*)-Non Infectious Dead (*D*)-Infectious Dead (*D*_*I*_)-Buried (*B*) compartmental model with corresponding viruses subpopulations (*V*_*k*_, *k* ∈ {*S, E, I, Q, H, T, R, D, D*_*I*_}) and free viruses in the environment (*V*_*F*_)

Let

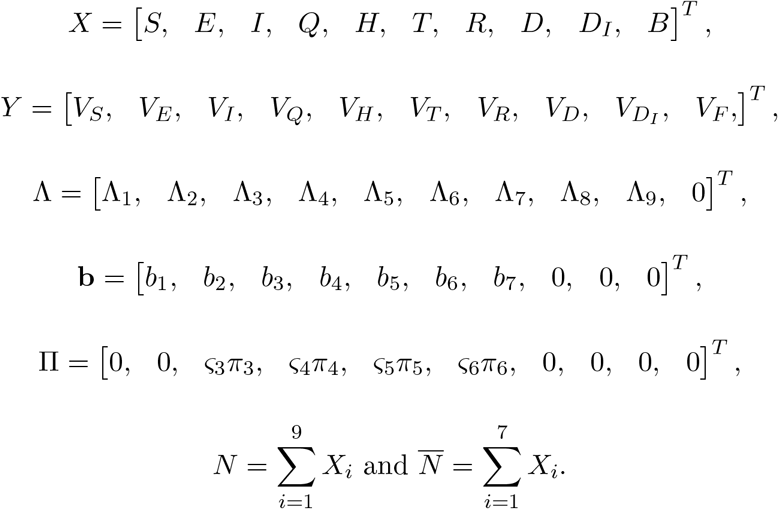

A mathematical model corresponding to the Figure 1 can be described as

- A mesoscopic model (intermediate scale of human gatherings):

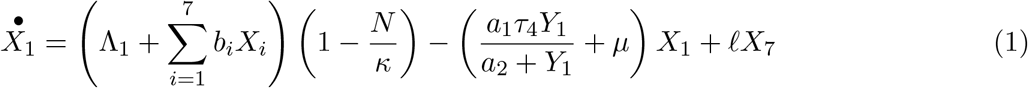

The first term in equation (1) corresponds to horizontal recruitment (immigration at rate Λ_1_) and vertical recruitment (birth at rates *b*_*i*_) in the susceptible human population. Population growth is restricted by logistic expression 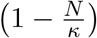. The limiting capacity *κ* models social distancing which reduces the number of individuals per unit of area. The second term represents the exposition and the natural death at rate *µ*. The last term corresponds to the lost of immunity at rate *𝓁*.

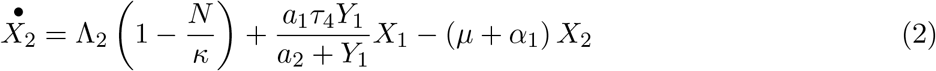

The first term in equation (2) corresponds to horizontal recruitment in the exposed human population. The second term represents the arrival of new exposed. The last term corresponds to the natural death at rate *µ* and the worsening of the disease at rate *α*_1_.

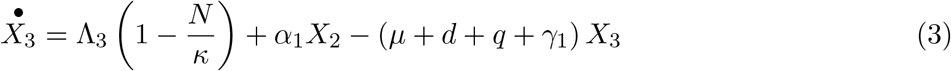

The first term in equation (3) corresponds to horizontal recruitment in the infectious human population. The second term represents the arrival of new infectious. The last term corresponds to the death at rate *µ* + *d*, the detection of the disease by screening test (at rate *q*) and the treatment at rate *γ*_1_.

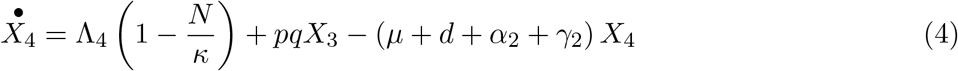

The first term in equation (4) corresponds to horizontal recruitment in the quarantined human population. The second term represents the arrival of new quarantined after testing an infectious at rate *pq*. The last term corresponds to the death at rate *µ* + *d*, the worsening of the disease (at rate *α*_2_ and the treatment at rate *γ*_2_).

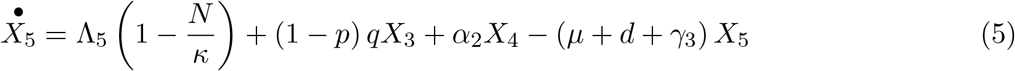

The first term in equation (5) corresponds to horizontal recruitment in the hospitalized human population. The second and third terms represent the arrival of new hospitalized after testing an infectious (at rate (1 − *p*) *q*) and disease worsening for quarantined (at rate *α*_2_). The last term corresponds to the death at rate *µ* + *d* and the treatment at rate *γ*_3_.

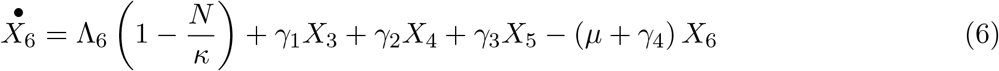

The first term in equation (6) corresponds to horizontal recruitment in the treated human population. The second, the third and the fourth terms represent the respective treating rates from the infectious (*γ*_1_), quarantined (*γ*_2_) and hospitalized (*γ*_3_). The last term corresponds to the death at rate *µ* and the recovering at rate *γ*_4_.

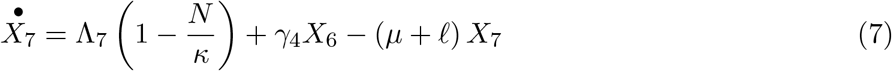

The first term in equation (7) corresponds to horizontal recruitment in the recovered human population. The second term represents the recovering rates of treated (*γ*_4_). The last term corresponds to the death at rate *µ* and the lost of immunity at rate *𝓁*.

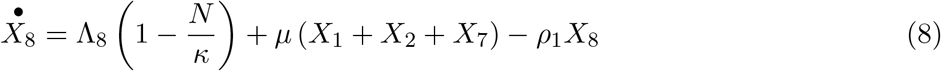

The first term in equation (8) corresponds to horizontal recruitment in the non-infectious dead human population. That recruitment could correspond to transportation or transfer of dead individuals for burial ceremonies like it is common in Cameroon or other valuable reasons. The second term represents the death of individuals in susceptible, exposed and treated compartment at rate *µ*. The last term corresponds to the burying at rate *ρ*_1_.

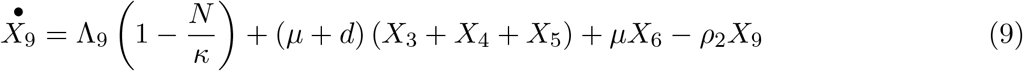

The first term in equation (9) corresponds to horizontal recruitment in the infectious dead human population. As in equation (8) that recruitment could correspond to transportation or transfer of dead individuals for burial ceremonies like it is common in Cameroon or other valuable reasons. The second term represents the death of individuals in infectious, quarantined, hospitalized at rate *µ* + *d*. The third term represents the death of individuals in recovered compartment *µ*. The last term corresponds to the burying at rate *ρ*_2_.

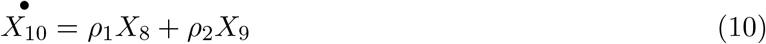

The first and the second terms in equation (10) correspond the burying of dead individuals at respective rates *ρ*_1_ and *ρ*_2_.

- And a microscopic model (virus scale):

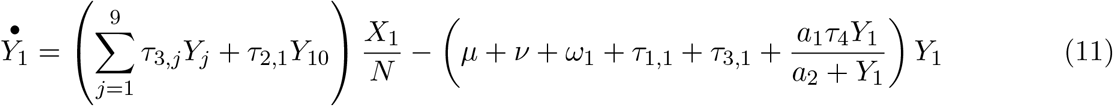

The first term in equation (11) represents the recruitment of viruses in susceptible population after a contact with environment (at rate *τ*_2,1_) and individuals of other compartments (at respective rates *τ*_3,*j*_). The second term corresponds to the disappearing of viruses by natural death (at rate *ν*), by the death of the human host (at rate *µ*), by disinfection measures applied by the host (at rate *ω*_1_), by the exchange of viruses with environment (at rate *τ*_1,1_) and other compartments (at rate *τ*_3,1_), and by the transition of the host from the susceptible compartment to the exposed compartment.

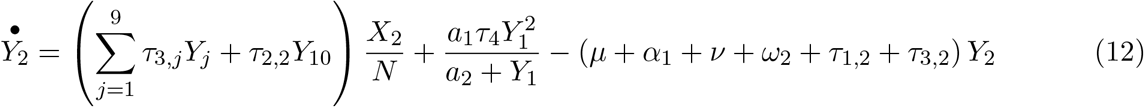

The first term in equation (12) represents the recruitment of viruses in exposed population after a contact with environment (at rate *τ*_2,2_) and individuals of other compartments (at respective rates *τ*_3,*j*_). The second term corresponds to the update in population of viruses due to the transition from susceptible status to exposed status at rate 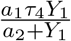. The third term represents the disappearing of viruses by natural death (at rate *ν*), by the death of the human host (at rate *µ*), by disinfection measures applied by the host (at rate *ω*_2_), by the exchange of viruses with environment (at rate *τ*_1,2_) and other compartments (at rate *τ*_3,2_), and by the transition of the host from the susceptible compartment to the exposed compartment at rate *α*_1_.

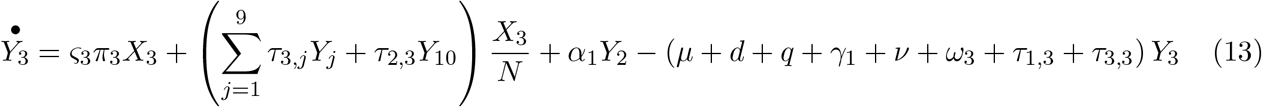

The first term in equation (13) represents the release of internal viruses by infectious individuals on themselves. The second term corresponds to the recruitment of viruses in infectious population after a contact with environment (at rate *τ*_2,3_) and individuals of other compartments (at respective rates *τ*_3,*j*_). The third term corresponds to the update in population of viruses due to the transition from exposed status to infected status at rate *α*_1_. The fourth term represents the disappearing of viruses by natural death (at rate *ν*), by the death of the human host (at rate *µ* + *d*), by disinfection measures applied by the host (at rate *ω*_3_), by the exchange of viruses with environment (at rate *τ*_1,3_) and other compartments (at rate *τ*_3,3_), by the transition of the host from the infectious compartment to the treated compartment at rate *γ*_1_, and by the transition of the host from the infectious compartment to the quarantined or the hospitalized compartment at screening rate *q*.

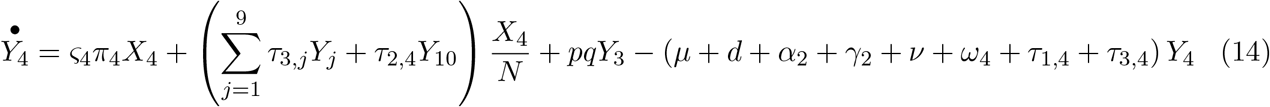

The first term in equation (14) represents the release of internal viruses by quarantined individuals on themselves. The second term corresponds to the recruitment of viruses in quarantined population after a contact with environment (at rate *τ*_2,4_) and individuals of other compartments (at respective rates *τ*_4,*j*_). The third term corresponds to the arrival of new quarantined from the infectious compartment at rate *pq*. The last term represents the disappearing of viruses by natural death (at rate *ν*), by the death of the human host (at rate *µ* + *d*), by disinfection measures applied by the host (at rate *ω*_4_), by the exchange of viruses with environment (at rate *τ*_1,4_) and other compartments (at rate *τ*_3,4_), by the transition of the host from the quarantined compartment to the hospitalized compartment at rate *α*_2_, and by the transition of the host from the quarantined compartment to the treated compartment at rate *γ*_2_.

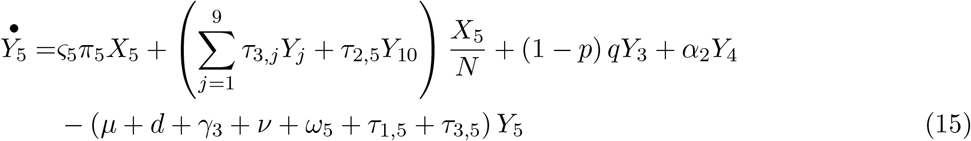

The first term in equation (15) represents the release of internal viruses by hospitalized individuals on themselves. The second term corresponds to the recruitment of viruses in hospitalized population after a contact with environment (at rate *τ*_2,5_) and individuals of other compartments (at respective rates *τ*_5,*j*_). The third term corresponds to the arrival of new hospitalized from the infectious compartment at rate (1 − *p*) *q*. The fourth term materializes the arrival of new hospitalized from the quarantined compartment at rate *α*_2_. The last term represents the disappearing of viruses by natural death (at rate *ν*), by the death of the human host (at rate *µ* + *d*), by disinfection measures applied by the host (at rate *ω*_5_), by the exchange of viruses with environment (at rate *τ*_1,5_) and other compartments (at rate *τ*_3,5_), and by the transition of the host from the hospitalized compartment to the treated compartment at rate *γ*_3_.

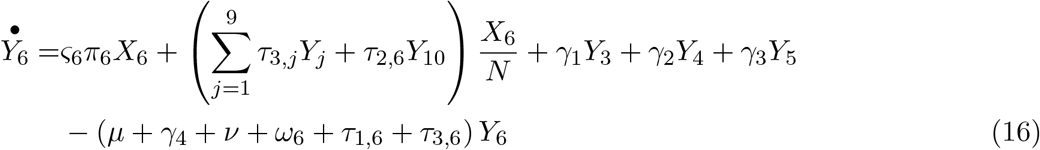

The first term in equation (16) represents the release of internal viruses by treated individuals on themselves. The second term corresponds to the recruitment of viruses in treated population after a contact with environment (at rate *τ*_2,6_) and individuals of other compartments (at respective rates *τ*_6,*j*_). The third, the fourth and the fifth terms correspond to the update in viruses population due to the transition from infectious, quarantined and hospitalized compartments respectively. The last term represents the disappearing of viruses by natural death (at rate *ν*), by the death of the human host (at rate *µ* + *d*), by disinfection measures applied by the host (at rate *ω*_6_), by the exchange of viruses with environment (at rate *τ*_1,6_) and other compartments (at rate *τ*_3,6_), and by the transition of the host from the treated compartment to the recovered compartment at rate *γ*_4_.

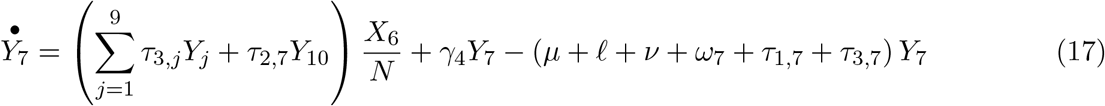

The first term in equation (17) corresponds to the recruitment of viruses in recovered population after a contact with environment (at rate *τ*_2,7_) and individuals of other compartments (at respective rates *τ*_7,*j*_). The second term corresponds to the update in viruses population due to the transition from treated compartment. The last term represents the disappearing of viruses by natural death (at rate *ν*), by the death of the human host (at rate *µ*), by disinfection measures applied by the host (at rate *ω*_7_), by the exchange of viruses with environment (at rate *τ*_1,7_) and other compartments (at rate *τ*_3,7_), and by the transition of the host from the recovered compartment to the susceptible compartment at rate *𝓁*.

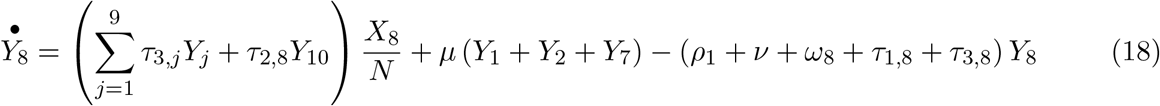

The first term in equation (18) corresponds to the recruitment of viruses in non-infectious dead population after a contact with environment (at rate *τ*_2,8_) and individuals of other compartments (at respective rates *τ*_8,*j*_). The second term corresponds to the update in viruses population due to the death of individuals belonging to susceptible, exposed and recovered compartments. The last term represents the disappearing of viruses by natural death (at rate *ν*), by the burial of the dead host (at rate *ρ*_1_), by disinfection measures applied on the dead host (at rate *ω*_8_), and by the exchange of viruses with environment (at rate *τ*_1,8_) and other compartments (at rate *τ*_3,8_).

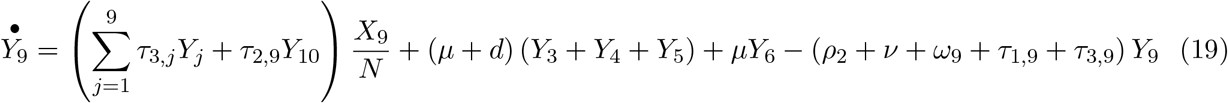

The first term in equation (19) corresponds to the recruitment of viruses in infectious dead population after a contact with environment (at rate *τ*_2,9_) and individuals of other compartments (at respective rates *τ*_9,*j*_). The second and the third terms correspond to the update in viruses population due to the death of individuals belonging to infectious, quarantined, hospitalized and treated compartments. The last term represents the disappearing of viruses by natural death (at rate *ν*), by the burial of the dead host (at rate *ρ*_2_), by disinfection measures applied on the dead host (at rate *ω*_9_), and by the exchange of viruses with environment (at rate *τ*_1,9_) and other compartments (at rate *τ*_3,9_).

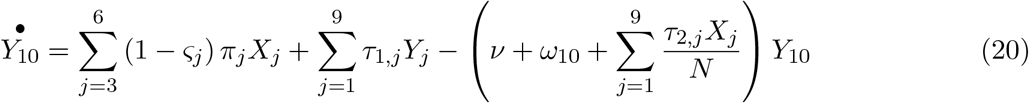

The first term in equation (20) corresponds to the release of virus in the environment by infectious, quarantined, hospitalized and treated populations. The second terms represents the recruitment of viruses in environment after a contact with individuals of each compartment (at respective rates *τ*_1,*j*_). The last term represents the disappearing of viruses by natural death (at rate *ν*), by disinfection measures (at rate *ω*_10_), and by the exchange of viruses from the environment to human hosts (at rate 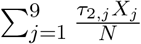).

The Monod^4^ type infection force 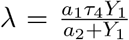 is the product of the mass action factor *a*_1_, the adequate contact probability 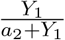 (the probability that viruses reach critical parts of the body like nose, mouth and eyes), the transfer rate *τ*_4_ and the susceptible body virus population size *Y*_1_. Although we focus explicitly on microscopic and mesoscopic scales, there is also a macroscopic scale of the global human population *X*_*G*_ =[*S*_*G*_ *E*_*G*_ *I*_*G*_ *Q*_*G*_ *H*_*G*_ *T*_*G*_ *R*_*G*_ *D*_*G*_ *D*_*I,G*_ *B*_*G*_] *T*. The equations at the macroscopic scale are very similar to those of the mesoscopic scale. Indeed, the limiting term *κ* can be replaced by +*∞* or higher value if a limiting capacity is still considered.

The parameters of the model (1)-(20) are described in Table 3.

The model (1) − (20) can be written in the form

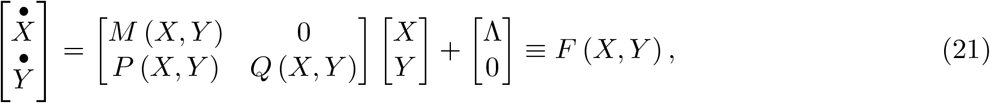

with *M, Q* Metzler and *P* having all its coefficients nonnegative.

#### Proposition 2.1

*Let* 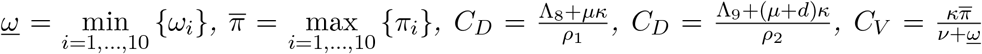 *and* 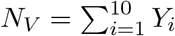

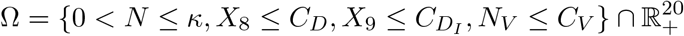

*is an attractor of the invariant positive orthant* 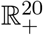 *with respect to the system* (21).

Proposition 2.1 shows the validity of the model (1) − (20). It shows the boundedness of the population density (*κ*) due to the social distancing. It also allows us to consider separately the microscopic dynamics (of viruses) and the macroscopic dynamics (of humans). Indeed, control strategies at the microscopic level concern the individual precautions to be applied (social distancing, mobility, disinfection of people and surfaces, mask wearing and treatment). At the macroscopic level, the global health policies are considered (social distancing, wearing mask, screening, rapid burying of infectious dead, isolation and treatment of infectious). Of course there are measures that act at both levels.

### 2.2 Equilibriums and asymptotic behaviors

Omitting the compartment of buried (*B* or *X*_10_), we can address the issue of existence of equilibriums. We are particularly interested in the ‘Disease Free Equilibrium’ (*DFE* = (*X*^*DFE*^, *Y* ^*DFE*^)) and other feasible’Endemic Equilibrium’ (*EE* = (*XEE, Y EE*)). It is clear that immigration in the infectious compartments with a constant positive rate makes impossible the existence of the DFE. Thus, we could suppose that Λ_*i*_ = 0, *i* = 2, *…*, 6, 8, 9. Such a hypothesis is not restrictive if the fight against the pandemic of corona virus is done globally (even Λ_*i*_ = 0, *i* = 1, 7) or a systematic control is applied at all the territorial frontiers (spatial isolation or lock-down). Notice that if the horizontal recruitment is proportional to the population with a rate less than the natural death rate then a disease free equilibrium is still possible (the population goes to extinction since death is dominant). If Λ_*i*_ = 0, *i* = 1, *…*, 9 then there is no horizontal recruitment and the only possibility of recruitment of new individuals is only from new births. That last consideration will be adopted here for simplicity.

#### Proposition 2.2

*Assume that* Λ_*i*_ = 0, *i* = 1, *…*, 9. *Then the following statements hold*.

i. *The trivial null equilibrium always exists* : *X* = *Y* = 0,
ii. *If b*_1_ > *µ, then the non-trivial disease free equilibrium (DFE) exists* :

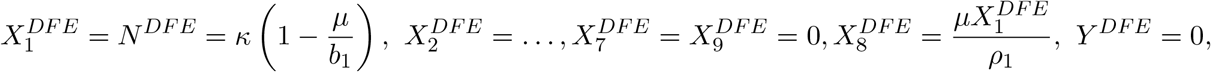
iii. *If b*_1_ > *µ and there is an endemic equilibrium (EE) then* 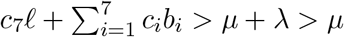 *where*

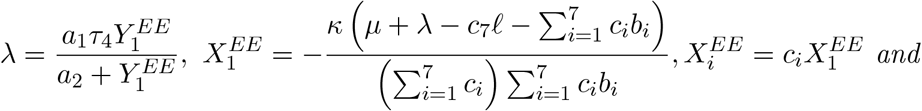

*the c*_*i*_*s are given as follows:*

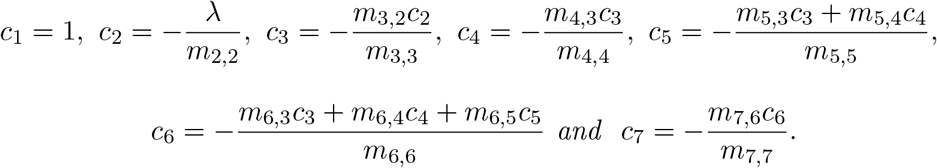

Let us consider the Jacobian matrix of (21) given by

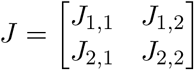

with

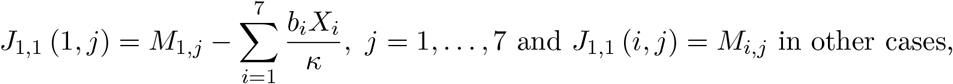

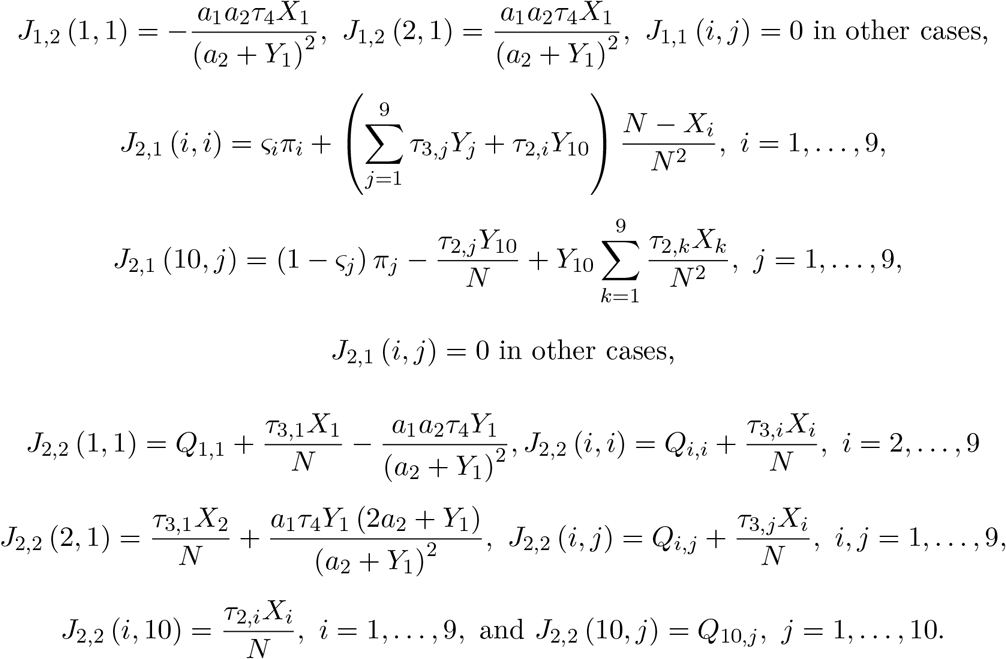

#### Proposition 2.3

*Assume that* Λ_*i*_ = 0, *i* = 1, *…*, 10.

i. *The null invariant set* {0}^9^ *×* ℝ_+_ *×* {0}^10^ *is locally asymptotically stable (LAS) if b*_1_ ≤ *µ*.
ii. *The null equilibrium X* = *Y* = 0 *is globally asymptotically stable (GAS) if b*_*i*_ ≤ *µ, i* = 1, *…*, 7.
iii. *The disease free equilibrium is locally asymptotically stable (LAS) if and only if b*_1_ > *µ and* 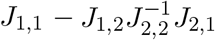 *is Metzler stable*.
iv. *If b*_1_ > *µ and λ* = 0 *then the disease free equilibrium for the macroscopic model (X* = *X*^*DF E*^, *Y* ∈ (ℝ_+_)^10^*) is GAS*.

Let *F* denote a 16 *×* 16 matrix given such as 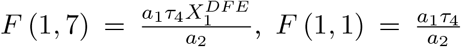 and *F* (*i, j*) = 0 otherwise. Let also *V* denote a 16 *×* 16 matrix given such as *V*_1,7_ = 0, *V*_*i,j*_ = *J* (*i* + 1, *j* + 1) if *i, j* = 1, *…*, 5, *V*_6,*j*_ = *J* (9, *j* + 1) if *j* = 1, *…*, 5, *V*_6,6_ = *J* (9, 9), *V*_6,*j*_ = *J* (9, *j* + 4) if *j* = 7, *…*, 16, *V*_*i*,6_ = *J* (*i* + 1, 9) if *i* = 1, *…*, 5, *V*_*i*,6_ = *J* (*i* + 4, 9) if *i* = 7, *…*, 16, *V*_*i,j*_ = *J* (*i* + 4, *j* + 4) if *i, j* = 7, *…*, 16, *V*_*i,j*_ = *J* (*i* + 1, *j* + 4) if *i* = 1, *…*, 5, *j* = 7, *…*, 16, (*i, j*) ≠ (1, 7) and *V*_*i,j*_ = *J* (*i* + 4, *j* + 1) if *i* = 7, *…*, 16, *j* = 1, *…*, 5. According to the Van Den Driessche and Watmough method in [56, 57, 58], the basic reproduction number *ℛ*_0_ is given by the spectral radius of the next generation matrix :

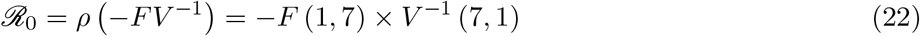

The effective reproduction number can also be computed by multiplying *ℛ*_0_ by the proportion of susceptible individuals [52]. *ℛ*_0_ can be biologically interpreted in this context as the average number of new viruses generated by an introduced virus during its lifespan.

The infection force *λ* plays an important role on the disease spreading. Subsequently, regarding the expression of *λ* in Table 3, we can see that the use of mask and avoiding to touch the face is crucial to limit disease spreading. The control *u* can be viewed as the fraction of time the mask is well used. *λ* can vary with time depending on individual human habits. It is interesting to depict how the disease behaves if *λ* is a decreasing function of time 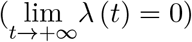.

#### Lemma 2.1

*Let us consider a linear differential system*

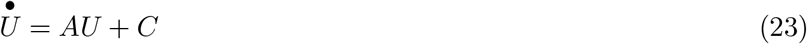

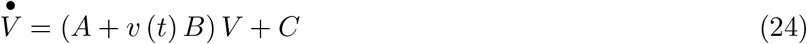

*where A is a constant n × n Metzler stable matrix, B is a constant n × n Metzler matrix, C is a constant n ×* 1 *matrix and v is a positive numeric function of the time. Assume that the solution* (*U, V*) *of* (23) *−* (24) *is bounded and* 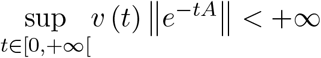. *Then* 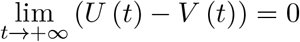. *This will happen in particular if* 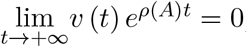, *where ρ* (*A*) *denotes the spectral radius of A*.

#### Theorem 2.1

*If b*_1_ *> µ and* 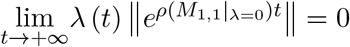 *then the DFE is GAS*.

Theorem 2.1 gives us a first way to limit disease spreading.

#### Theorem 2.2

*Assume that* Λ_*i*_ = 0, *i* = 1, *…*, 10 *and b*_1_ *> µ. Let D denotes a diagonal matrix such that D*_*i,i*_*is the sum of absolute values of only negative terms in the expression of* 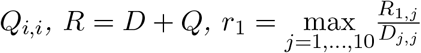 *and* 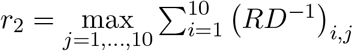. *Then the DFE is GAS if*

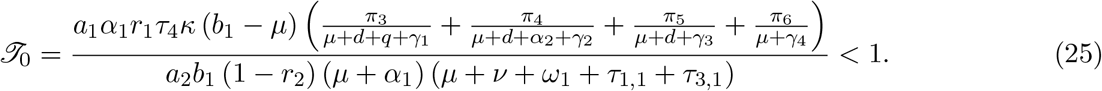

Theorem 2.2 provides a sufficient condition that ensures the stability of the disease free equilibrium. Moreover, it shows that avoiding to touch the face and the frequency of wearing mask by susceptible population (*u*_1_ via *τ*_4_), the mobility (*m* via different contact rate among people *τ*_3_, and with the environment *τ*_1_), the disinfection (washing of hands included in) by susceptible population (*ω*_1_) and social distancing (reducing *κ*) are crucial parameters in order to stop the spreading of the disease. The biological interpretation of *𝒯*_0_ is the following. During its lifespan 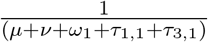 a virus infects a susceptible individual at the adequate rate 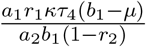. Once the susceptible individual is infected he becomes infectious with a probability 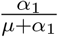 and will produce 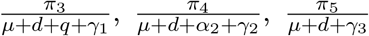 and 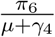 viruses during its respective stays in compartments of infectious, quarantined, hospitalized and treated.

By setting

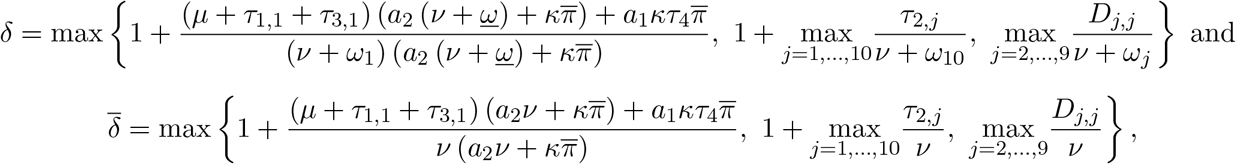

we can see using the boundedness of *N*_*V*_, that 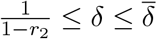. Hence, Theorem 2.2 also permits us to evaluate given *κ, u* and *m* the minimum frequency at which the disinfection should occur. For example, if 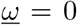 probably due to the management of death persons, then the condition *𝒯*_0_ ≤ 1 is satisfied when

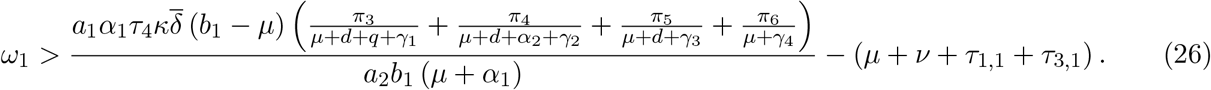

Notice that by disinfection we mean a global action on the body, so it would be necessary to evaluate the specific contribution of hands washing.

Figure 2 is an illustration of the asymptotic behavior of cumulative infectious individuals depending on the relative position of *𝒯*_0_ with respect to 1. The smaller *𝒯*_0_ is, the smaller is the amplitude of the peak and the earlier it occurs. If the mask is never adopted then the disease prevalence can pass above 30% and stabilize around 3.5%.

**Figure 2.**
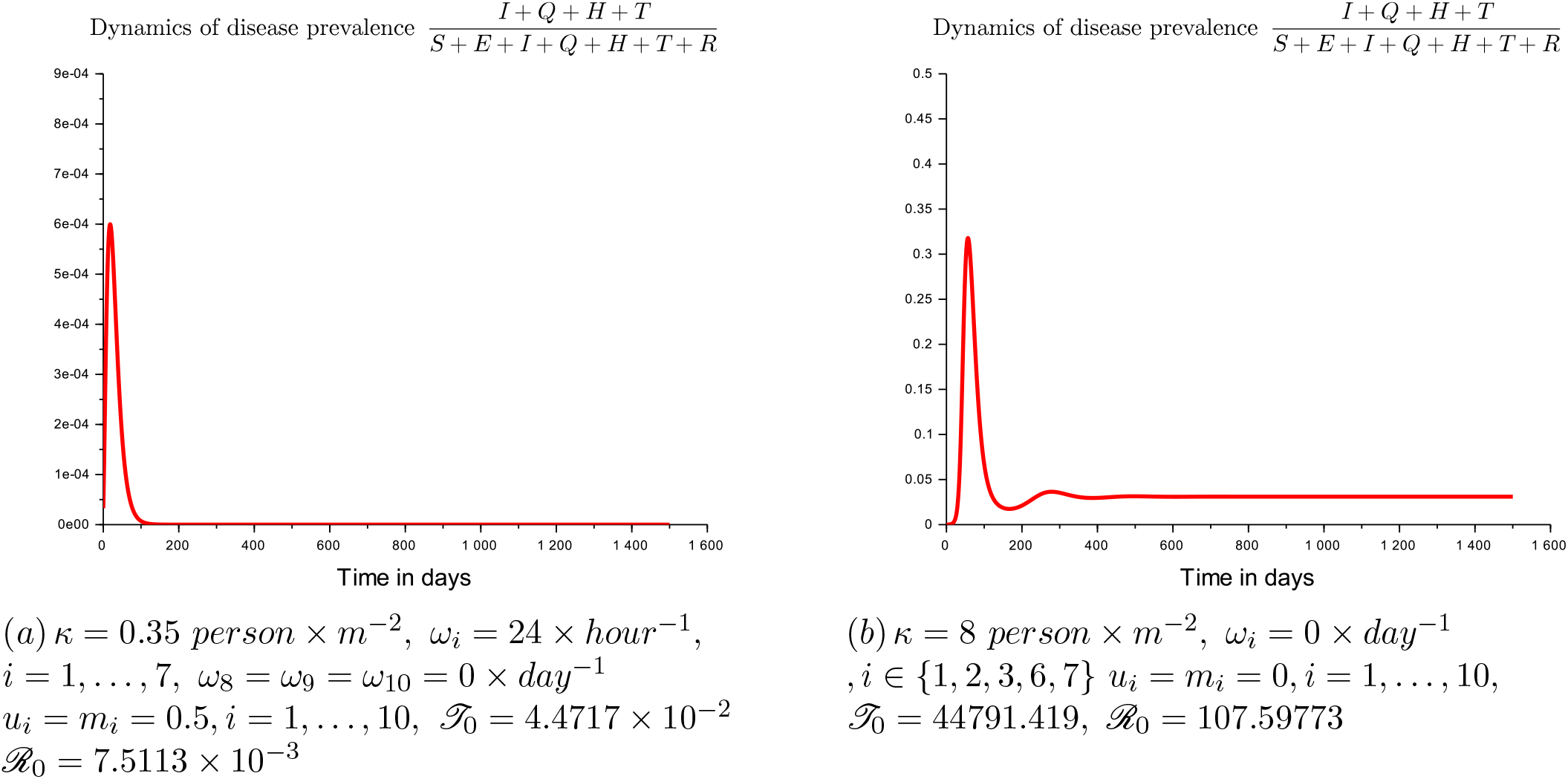
Asymptotic behavior of infectious dynamics depending on *𝒯*_0_

We end the section with a theorem giving an upper bound to the time of extinction of the disease.

#### Theorem 2.3

*Let* 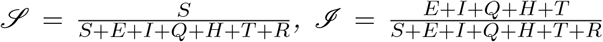 *and 𝒜 denotes the effective area of the spatial study zone in m*^*−*2^ *(ie the area of the zone each individual is allowed to move in during the lock-down). Assume that ∀t* ≥ 0, *λ* (*t*) = *λ*_0_*e*^*−αt*^, *α >* 0. *Then*

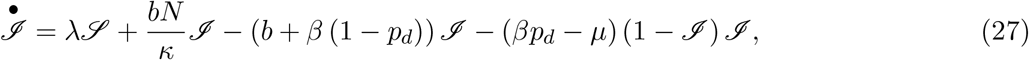

*where* 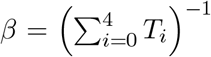, *p is the probability to die during the stay in infected or infectious compartments (E, I, Q, H, T) and the T*_*i*_*s denote the average times spent in those compartments. Moreover, if σ*_1_ = min {*α, β* (1 *− p*_*d*_)} *then*

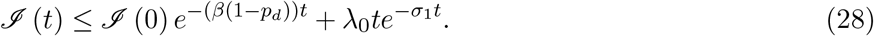

*Furthermore, if* 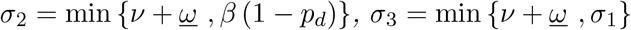 *then*

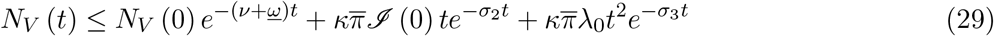

*and the respective first times* 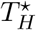 *and* 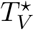 *such that respectively* 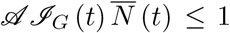*and 𝒜 N*_*V*_ (*t*) ≤ 1 *satisfy*

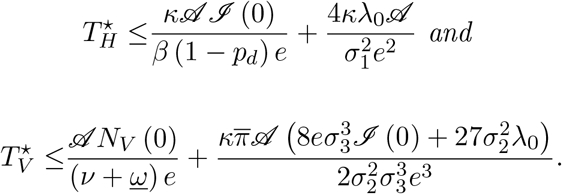

As we can see in the expressions of 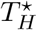 and 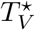 that the higher *κ, λ*_0_, *ℐ* (0), *N*_*V*_ (0) and *𝒜* are, the longer the time of disease extinction 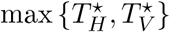 is. If the lock-down is applied, then the effective global population to consider is 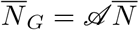 which represents the number of alive persons admitted in a given closed area. Hence, the lock-down level can be expressed by the ratio 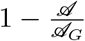, where *𝒜*_*G*_ denotes the real area of the physical study zone (town, district, department, region, country, continent, or the whole world).

## 3 A focus on Cameroonian context

### 3.1 Estimation of parameters

There are several approaches for the estimation of the parameters of a model. A first method is to discretize the differential equation model with respect to available time series and to fit unknown parameters by reducing the distance between the numerical solution and observed time series [7]. Another method consists of empirical estimating function linking the state of the system with the time using times series and after in estimation through the considered model [6, 16, 40, 47].

Let *CRI* (*t*), *CRQ* (*t*), *CRH* (*t*), *CRR* (*t*) and *CRD* (*t*) respectively denote the cumulative numbers of reported infectious cases, of reported quarantined, of reported hospitalized (active cases), of reported recovery, and of reported dead at time *t* ≥ *t*_0_, where *t*_0_ denotes the date of the first introduction of an infectious to be evaluated. Notice that those values will be divided by the cumulative surface of living areas to match with our model. We have

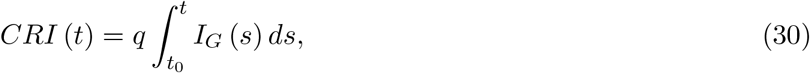

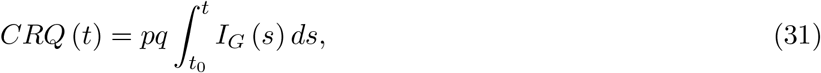

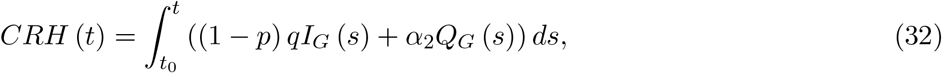

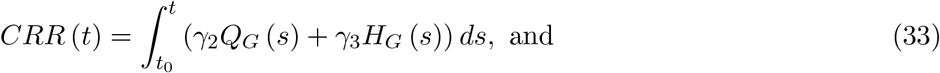

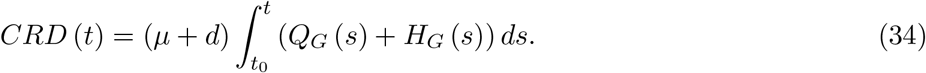

According to some cultural practices in Cameroon, we will assume that the average time before burying a dead person which is not quarantined is 3 days 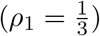 while the burying immediately follows the death of an infectious (*ρ*_2_ = 1). From the literature we will assume that 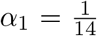 and 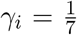, *i* = 1, *…*, 4. It is not clearly established in the literature that there is a real immunity, but according to [32], we will suppose that *ℒ* = 2.74 *×* 10^*−*3^. The recommended social distancing parameter *κ* in Cameroon is 1 *person × m*^*−*2^ but we will estimate the real value of *κ* according to available data and our model.

Regarding the fast growth of reported infectious cases, we assume that *CRI* can be approximated through the following regression model

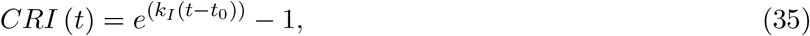

We also assume that 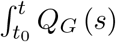 and 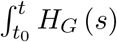 *H*_*G*_ (*s*) *ds* can be approximated through similar log-linear models. We adopt the following estimation procedure :

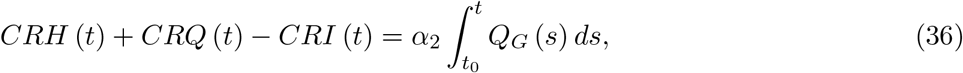

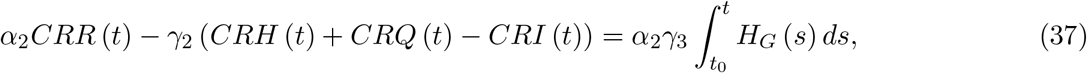

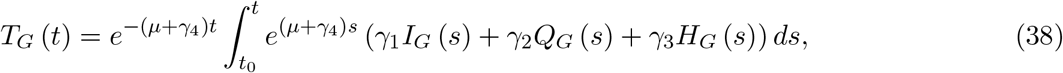

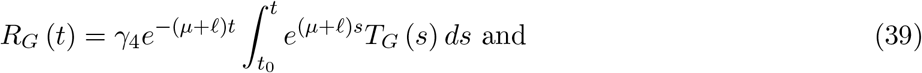

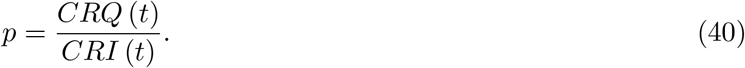

If *t*_1_ denotes the smallest solution to the equation *Q* (*t*) = 1 then it appears natural to estimate *q* by the relation

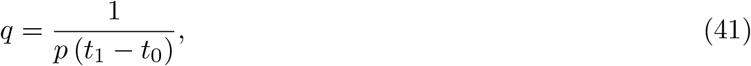

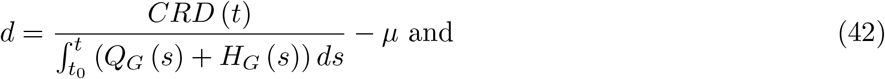

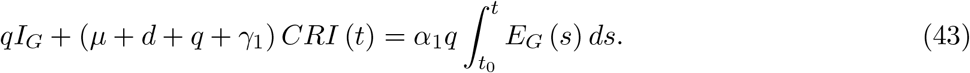

It appears difficult with available data to estimate explicitly 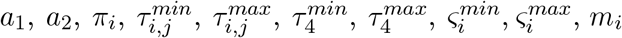 and *u*_*i*_. Regarding public transport conditions in Cameroon the maximum value of *κ* can be fixed to 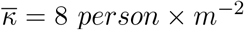. According to the density of population per unit of surface^5^ the minimum value of *κ* is fixed to *<u>κ</u>* = 5.003 × 10^−5^ *person × m*^*−*2^. Without social distancing at the disease outbreak, we have 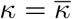 and the infection term *λS* is proportional to the global population (say *S*_*G*_) due to the mass action. *S* being bounded from above by 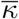 according to the model, we have 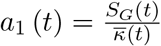. Notice that *<u>κ</u>* and 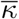 are non decreasing functions of *S*_*G*_. Thus, we can simplify by assuming that *a*_1_ is definitely a constant given by 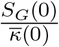. It is possible to estimate *S*_*G*_ and *N*_*G*_ (global population of alive or dead except buried individuals) similarly to *T*_*G*_ and *R*_*G*_. Under the hypothesis of homogeneity of the population the values of *S, E* and *I* can be estimated by multiplying the respective proportions in the global population by the threshold 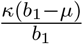 (for example 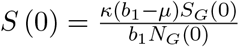). The term 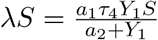 can be estimated using the relations

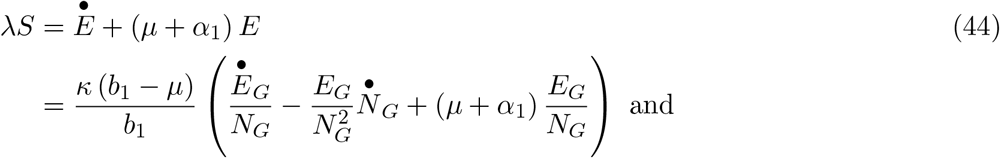

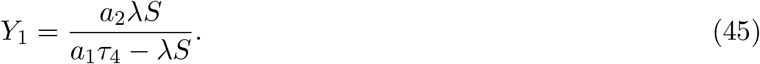

We used the daily data available on the website www.data.gouv.fr for our estimates. We also used the situation reports on Covid-19 in Cameroon available on the website https://www.humanitarianresponse.info. The estimated parameters are given into the Table 4. Figure 3 shows the cumulative reported infectious cases in Cameroon from the 2^*nd*^ of March to the 07^*th*^ of June 2020.

**Figure 3.**
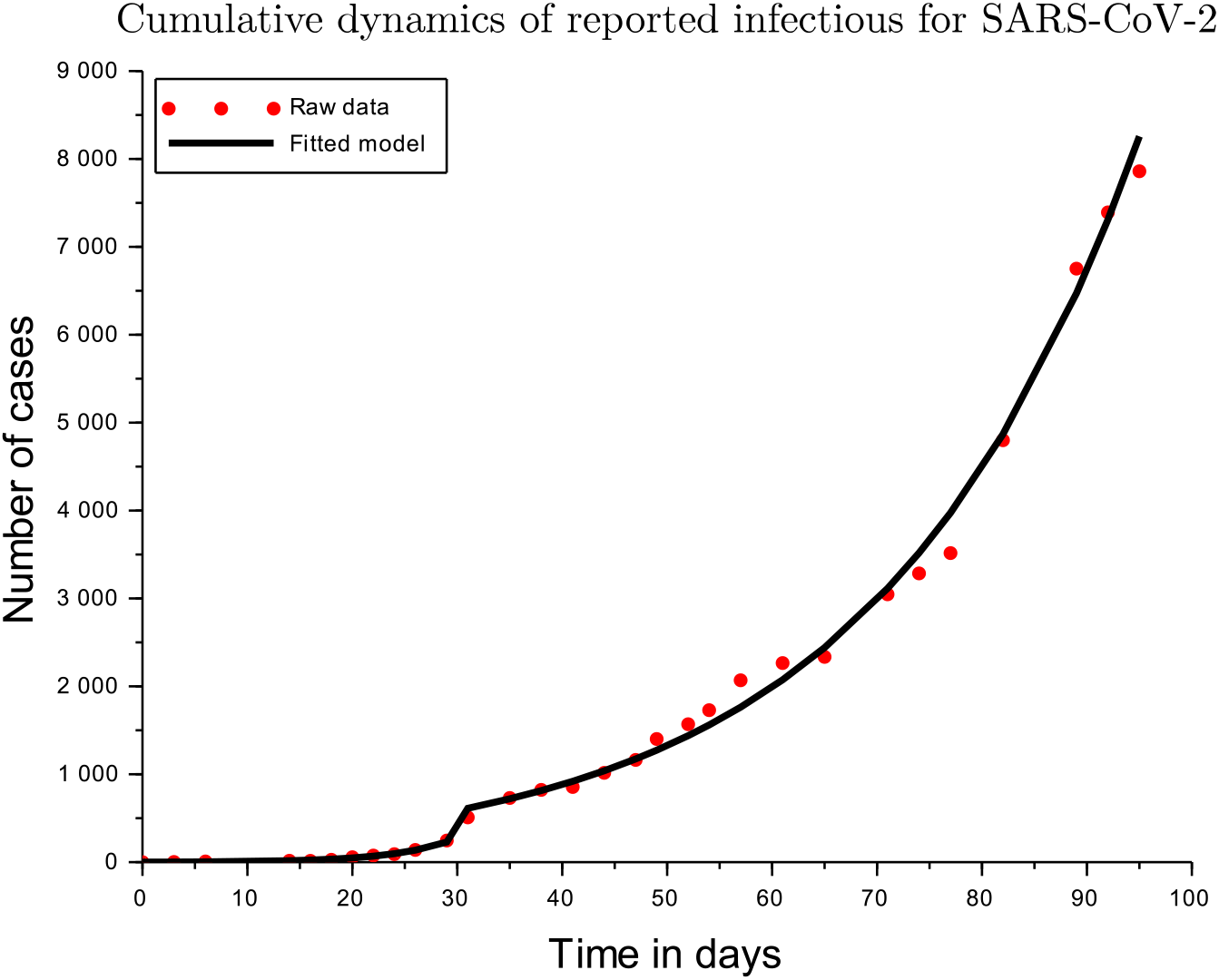
Cumulative reported infectious from the 2^*nd*^ of March to the 07^*th*^ of June 2020.

According to the values in Table 4, relation (26) and Theorem 2.2, if people never wear a mask, no social distancing is applied and only susceptible individuals disinfect themselves, then the rate of **complete** disinfection (*ω*_1_) should not be less than 20 *day*^*−*1^ (ie about once every 72 minutes) in order to have *𝒯*_0_ = 7.3163071 and *ℛ*_0_ = 0.887382. If at least the social distancing of 1 *person × m*^*−*2^ is respected then the lower bound value of *ω*_1_ drops to 3 *day*^*−*1^ (ie about once every 30 minutes) in order to have *𝒯*_0_ = 5.7754491 and ℛ_0_ = 0.7064687. Thus, a complementary application of all mitigation measures is mandatory.

### 3.2 Effects of controls on disease dynamics

This section aims at evaluating first the sensitivity of different control parameters with respect to the thresholds *ℛ*_0_ and *𝒯*_0_. The second fold of this section is to survey the effect of some control parameters on the asymptotic behavior of the disease dynamics. Following Theorem 2.1, we define the convergence index

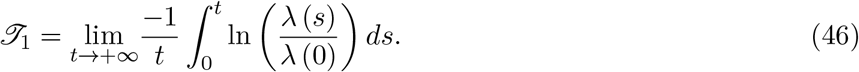

*𝒯*_1_ is an indicator of the rate of the potential convergence of the disease dynamics towards an equilibrium. Indeed, if *𝒯*_1_ *>* 0 then there is an exponential convergence to the disease free equilibrium. If *𝒯;*_1_ ≤ 0 then there is an endemicity of the disease, in particular if *𝒯*_1_ is near to zero then we are in the neighborhood of an endemic equilibrium. *𝒯*_1_ is an estimation of the key parameter *α* which permits according to Theorem 2.3, to bound the average time the disease is expected to disappear.

There are several methods for sensitivity analysis in the literature (see [11, 42] for example). The normalized forward sensitivity index (of variable *u* with respect to parameter *p*) 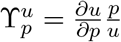 defined in [11] provides an idea on effect of a local perturbations around a given situation in terms of the values of the parameter *p*. Paper [42] presents several alternative methods of assessing the global effect of variation on coefficients: Pearson correlation coefficient (CC), Spearman rank correlation coefficient (RCC), partial correlation coefficient (PCC), standardized correlation coefficient (SCC), standardized rank correlation coefficient (SRCC), partial rank correlation coefficient (PRCC) and Extended Fourier amplitude sensitivity test (eFAST). The authors in [42] recommended PRCC and eFAST as robust sensitivity indexes to be used respectively in cases of monotonic and non-monotonic sensitivities. These indexes are based on samples obtained through latin hyper cube sampling, Monte-Carlo sampling or any other appropriate experimental design [26]. Hence, there are statistics and confidence intervals or region can be provided in order to decide on their significance.

We adopt the PRCC with a three levels (*−*1, 0, +1) fractional experimental design with the factors *κ, q, m*_*i*_, *u*_*i*_ and *ω*_*i*_, *i* = 1, *…*, 10. Such an experimental design allows us to get a statistical approximation of optimal parameters based on a quadratic model obtained by regression. Ideally, strategies should be specific to different compartments. However, since it is difficult to know the status of people, it is reasonable to apply the same strategies for “similar” compartments. Thus, we reduce the factors by considering the classes [1] *≡* {1, 2, 3, 6, 7}, [4] *≡* {4, 5}, and [8] *≡* {8, 9, 10}. We also assume that *u*_8_ = *u*_9_ = *u*_10_ = 0. Finally, we have 10 factors with two responses *𝒯*_0_ and *𝒯*_1_. We consider a 3 fractional experimental design with a sample size 81. The experimental matrix is computed using Box calculus for the aliases with second order interactions. Table 1 provides us with the codification of factors values that have been arbitrarily chosen but appear realistic.

**Table 1:** Codification of factors values

| Parameter | Level -1 | Level 0 | Level +1 |
| --- | --- | --- | --- |
| $\kappa$ | 1 | 4.5 | 8 |
| $q$ | 0 | 24 | 48 |
| $u_i$ | 0 | $5 \times 10^{-1}$ | $9.99 \times 10^{-1}$ |
| $m_i$ | 0 | $5 \times 10^{-1}$ | 1 |
| $\omega_i$ | 0 | 24 | 48 |

Figure 4 shows graphically the effects of control parameters on *ℛ*_0_, *𝒯*_0_ and *𝒯*_1_. *u*_1_, *m*_1_, *ω*_1_ and *ω*_10_ are in decreasing order the more sensitive parameters with respect to *ℛ*_0_. Thus, an efficient control strategy should prioritize the usage of mask, the reduction of mobility and the disinfection in class [1] compartment, and the disinfection of the environment. *u*_1_, *ω*_1_, *ω*_4_ and *ω*_10_ are in decreasing order the more sensitive parameters with respect to *𝒯*_0_. Thus, an efficient control strategy based on *𝒯*_0_ should prioritize the usage of mask in class [1], and disinfection in classes [1], [2] and the environment. *m*_1_, *u*_1_, *ω*_1_, and *ω*_10_ are in decreasing order the more sensitive parameters with respect to *𝒯*_1_. Thus, to accelerate the convergence toward the disease free equilibrium the fighting policies should prioritize the reduction of mobility in class [1], the frequent usage of mask, the disinfection of people in class [1] and the disinfection of the environment.

**Figure 4.**
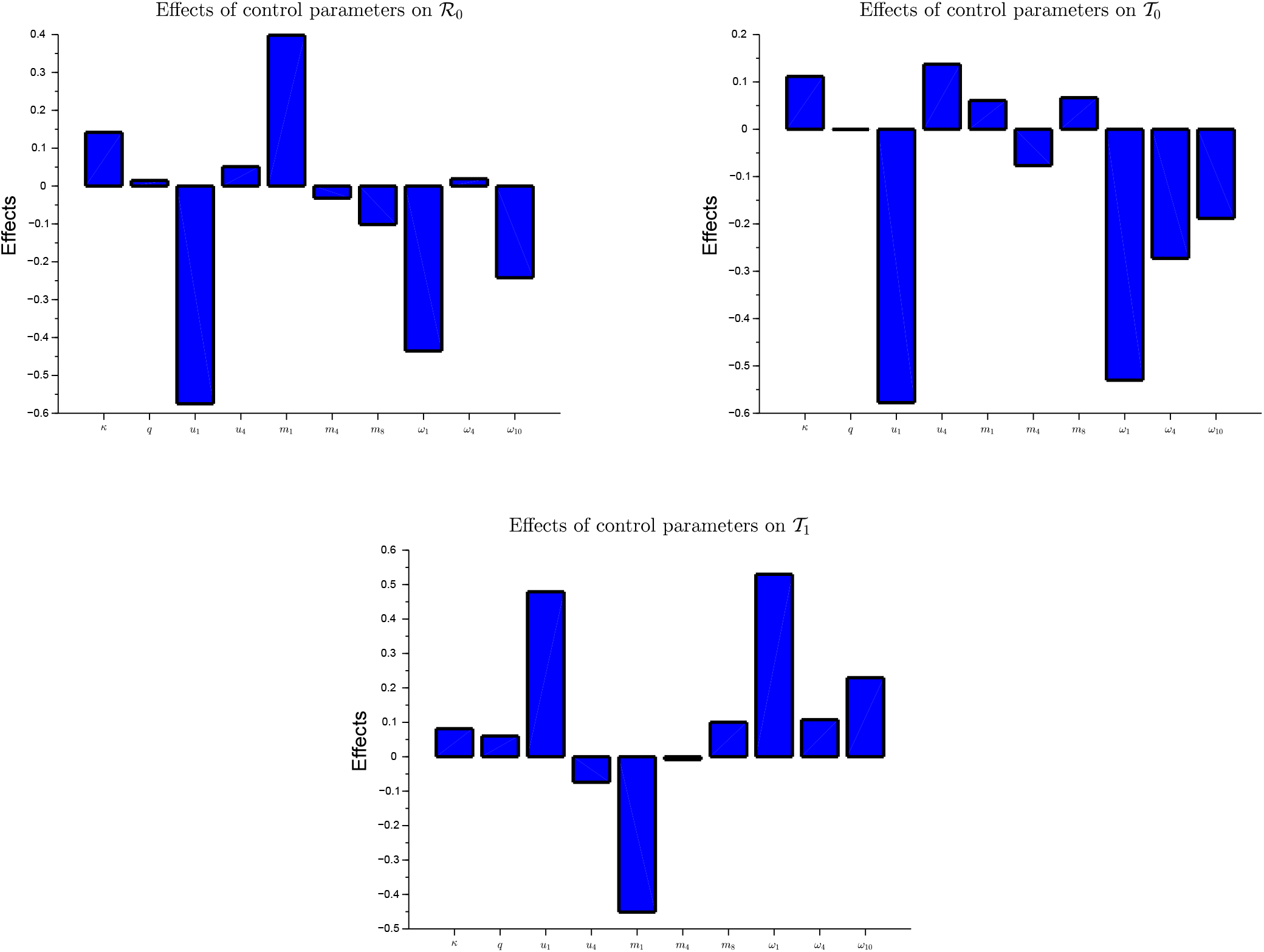
Effects of control parameters on *R*_0_, *T*_0_ and *T*_1_

Globally, the usage of mask in class [1] compartment is the most sensitive control followed by the reduction of mobility and the disinfecting in class [1], and the disinfection of the environment. As we can observe, the effects of the social distancing and the screening, the effect of using masks in class [4], and the mobility in classes [4] and [8] are not very significant. However, as we will observe in section 4, those controls have influence on the economical point of view.

## 4 An optimal control analysis

### 4.1 Design of the control strategy

The control parameters we consider here are mainly the limit density of population *κ*, the probability of mobility *m*, the proportion of time spent wearing a mask *u*, the disinfection rate *ω* and the reporting rate of infectious (through screening test especially) *q*. Indeed, those controls appear explicitly in the expression of the threshold *𝒯*_0_ which will play a similar role to the basic reproduction number. Since the threshold *𝒯*_0_ appears important for the asymptotic behavior of the disease, it could be interesting to minimize an analog to the effective reproduction number : 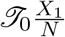. Unfortunately, the expression of *𝒯*_0_ is very complex and it is not differentiable with respect to all its parameters. This irregularity involves an additional complexity in the optimization procedure. Thus, we will consider the infection rate *λX*_1_ instead of 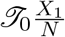. *λX*_1_ takes into consideration the dynamics of viruses.

The medical monitoring of a patient during his infectious period has an economical cost *c* that can be evaluated depending on the time spent before recovery and on the expenses during each stay in a disease compartment. The expression of *c* should depend on *c*_*g*_, the daily revenue of a person in health which will be lost due to the disease. The application of each control involves an economical cost proportional to the size of the population and to the time. We will note the cost per unit of time and per individual *c*_*u*_ for *u* and *c*_*ω*_ for *ω*. Reducing *m* and *κ* involves a relative reduction in economical activities and subsequently a reduction of gross domestic product assumed proportional to *mκ* (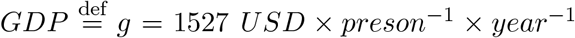 in 2018^6^). A screening test has a cost *c*_*s*_ which is not clearly defined according to the media. We will assume that *c*_*s*_ = 10 *USD* in average. Since it is difficult to really estimate unreported infectious, it seems better to define general measures to be applied by everyone independently of the status. Thus, we will assume here that *τ*_1,*i*_ = *τ*_1_, *τ*_2,*i*_ = *τ*_2_, *τ*_3,*i*_ = *τ*_3_, *u*_*i*_ = *u, ω*_*i*_ = *ω, ς*_*i*_ = *ς, m*_*i*_ = *m*. Let *C* = (*κ, q, u, m, ω*). We consider the following functional cost per area *𝒜* :

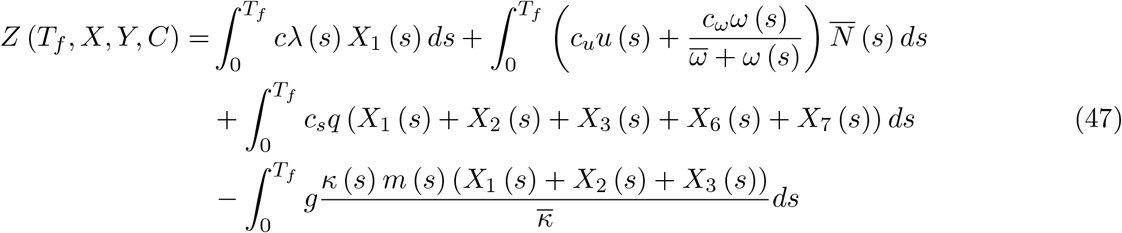

where 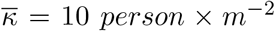 denotes the usual average density of population per unit of surface, and 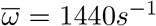 is the rate of hands washing corresponding to spending the half of time for disinfection. The negative term in the expression of *Z* represents the reduction of economic losses when people are free to move around in their various economic activities. A negative value of the cost means that globally the losses due to Covid-19 are not greater than the production of wealth.

We assume that an efficient hands washing takes about 30*s*. Thus, if *ω*^*−*1^ = 60*s* the time spent hands washing is half of the time between two consecutive washes. On the other hand, when *ω* tends to +*∞* then the proportion of time spent to wash hands is 1. A function of *ω* displaying the behavior explained above is 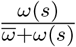 with 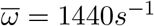. We suppose that the disinfection costs is *c*_*ω*_ = 1.736 *×* 10^*−*5^ *USD × person*^*−*1^ *× s*^*−*1^ in average. We also assume that after using a mask for a given time (4 hours) it is systematically changed for a new mask having a certain unitary cost (1 *USD*). Hence, *c*_*u*_ = 0.25 *USD × person*^*−*1^ *× hour*^*−*1^.

Let *T*_*i−*2_ and *c*_*i−*2_ denote respectively the time spent by an individual in the compartment *i* = 2, *…*, 6 and the cost generated by unit of time. According to probabilistic properties of Poisson processes [8, 17, 33], we have

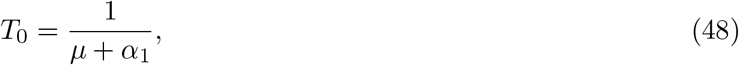

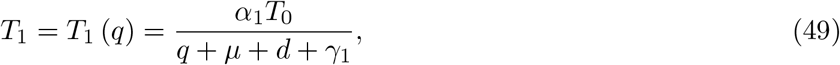

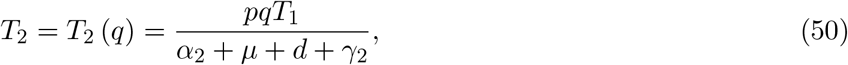

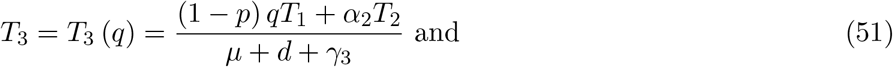

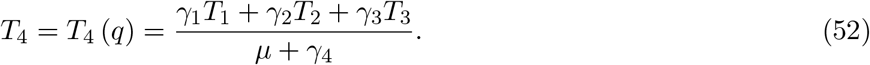

One can associate a cost *c*_*d*_ per person which is related to his burial ceremony. The probability to die during the stay in disease compartments *I, Q* and *H* is given by

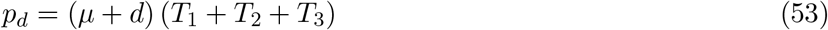

Hence,

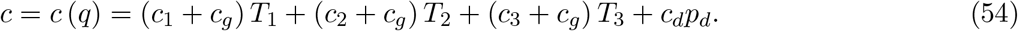

Table 2 summarizes the different unit costs we mentioned above.

**Table 2:**
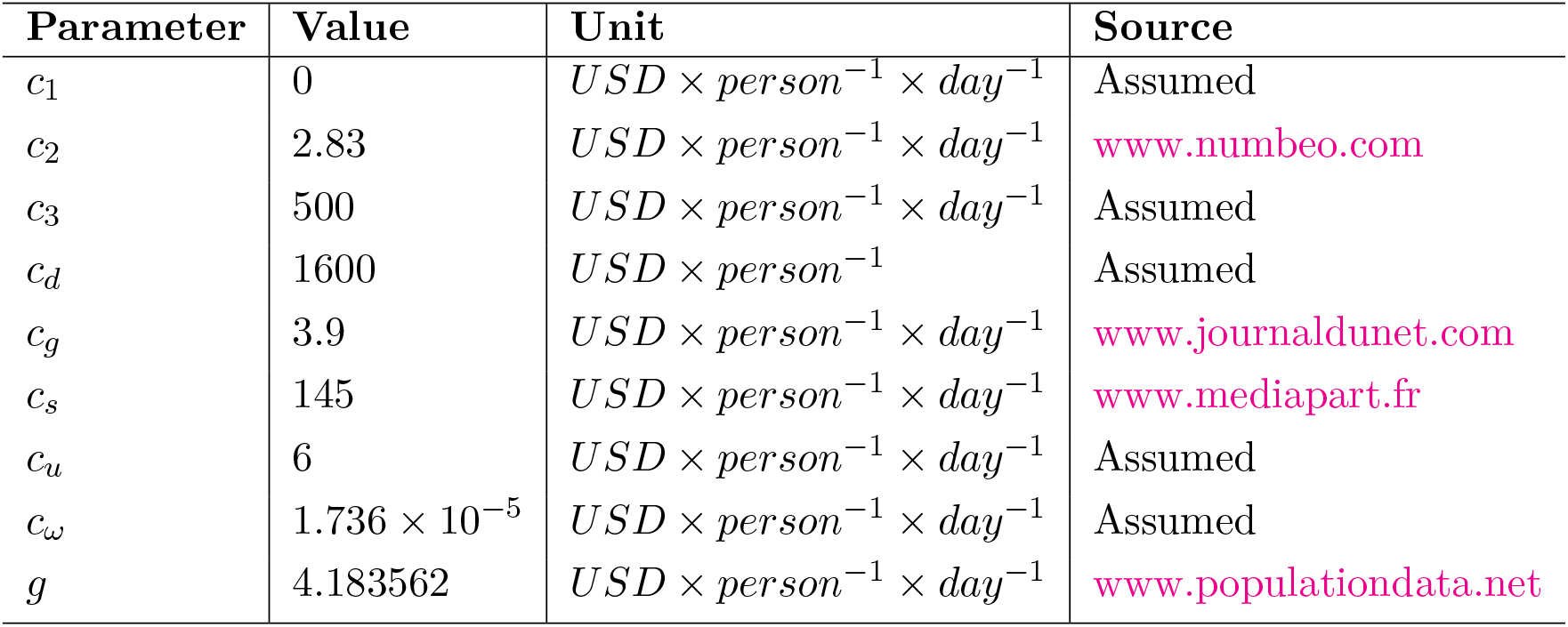
Baseline values of elementary costs

**Table 3:**
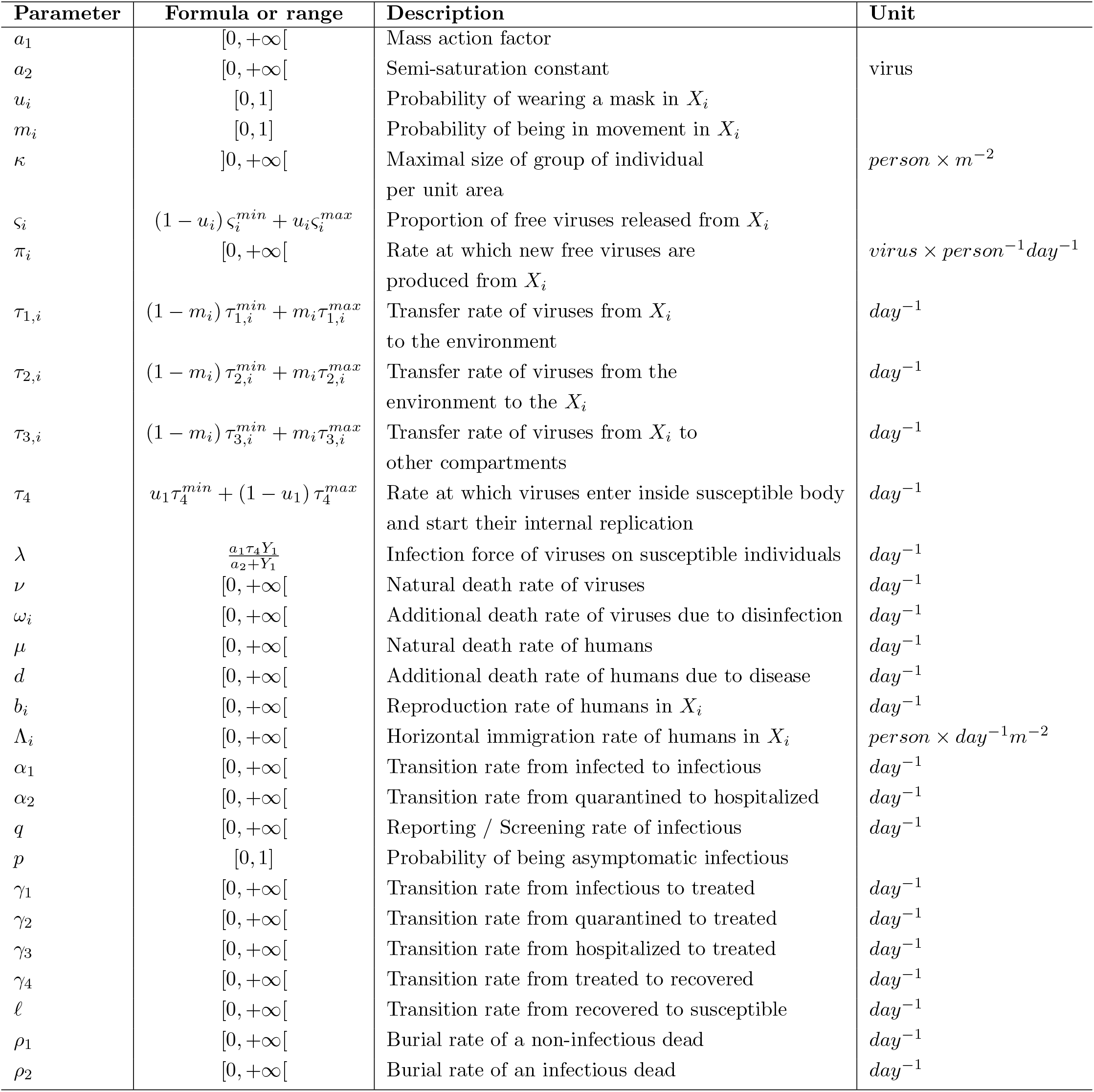
Description of the parameters of the model (21)

Let us consider the Hamiltonian

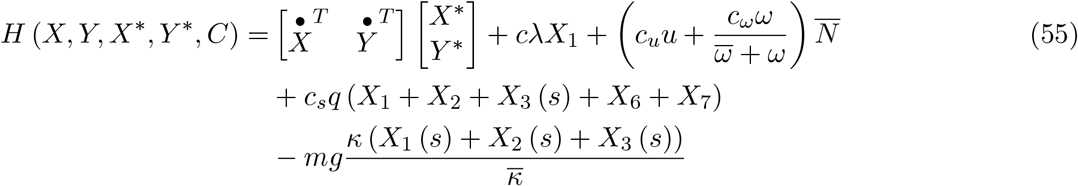

where the adjoint state 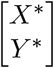 satisfies

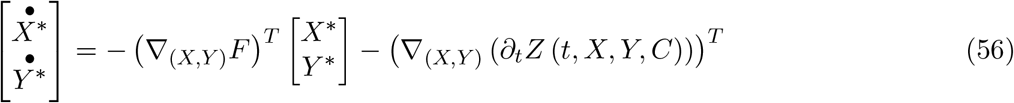

and

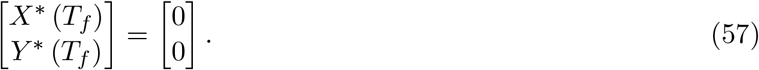

The optimal control *C*^*∗*^ will be given by the optimality condition

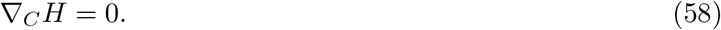

Notice that *C*^*∗*^ is projected into the admissible set 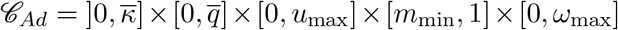 where 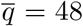 (persons are tested in average every 30 minutes), *u*_max_ = 0.9, *m*_min_ = 0, *ω*_max_ = 24. The existence of a unique optimal strategy is guaranteed by the fact that the strict convexity of the Hamiltonian with respect to *C* and the regularity of the optimal feedback strategy *C*^*∗*^ (*X, Y, X*^*∗*^, *Y* ^*∗*^). For more details on the determination of optimal control we refer to the books [2, 36] and references therein.

#### Theorem 4.1

*There is a unique piecewise continuous optimal control C*^*∗*^ *for the model* (1)*-*(20) *characterized by Algorithm 1*.

### 4.2 Numerical simulations and discussion

In order to numerically determine the optimal control strategy, we implement the forward-backward sweep method described in Chapter 3, page 101 of [2]. It is an iterative approach that consists of replacing the feedback by a guess value first, solving the forward state problem, solving the backward adjoint problem and then updating the feedback using the gradient direction with a step length obtained by the golden section method. The algorithm stops when the updates of the control does not significantly change its values according to a chosen threshold. That method is similar to the one given into the Chapter 3 of [36], but it permits to better control the convergence. Notice that one could also envisage a method which minimizes the distance between the final condition of the adjoint problem obtained for a guess initial condition and the reference final condition. That method could be a multidimensional variant of the secant algorithm [18, 30].

We found after several simulations that, if we globally consider the economic losses and gains then, no specific control effort is needed additionally to take care of hospitalized patients and putting in quarantine of infectious reported individuals as it is already done. Probably that is a consequence of the high recovery rate of patients and the relatively small death probability. However, since the logistic resources for quarantine and hospitalization are limited, the economical losses in terms of gross domestic product (due to social distancing, reduction of mobility and lock-down) can be penalized in the cost functional *Z*. That is done in the rest of the section.

Figure 5 is an illustration of the optimal control strategy against Covid-19 and the evolution of the infection force *λ*. We can observe that social distancing and disinfection frequency are the main elements of the optimal control strategy. Social distancing permits to have no restriction on mobility. The use of mask is useful but not necessary when social distancing and regular disinfection are respected. Notice that if the mask is misused it is useless, and it can promote other respiratory complications. As the sensitivity analysis showed the screening has no significant effect on disease behavior and it is economically better to keep *q* at its minimal value corresponding to self decision to be tested or the hospitalization of severe disease cases.

**Figure 5.**
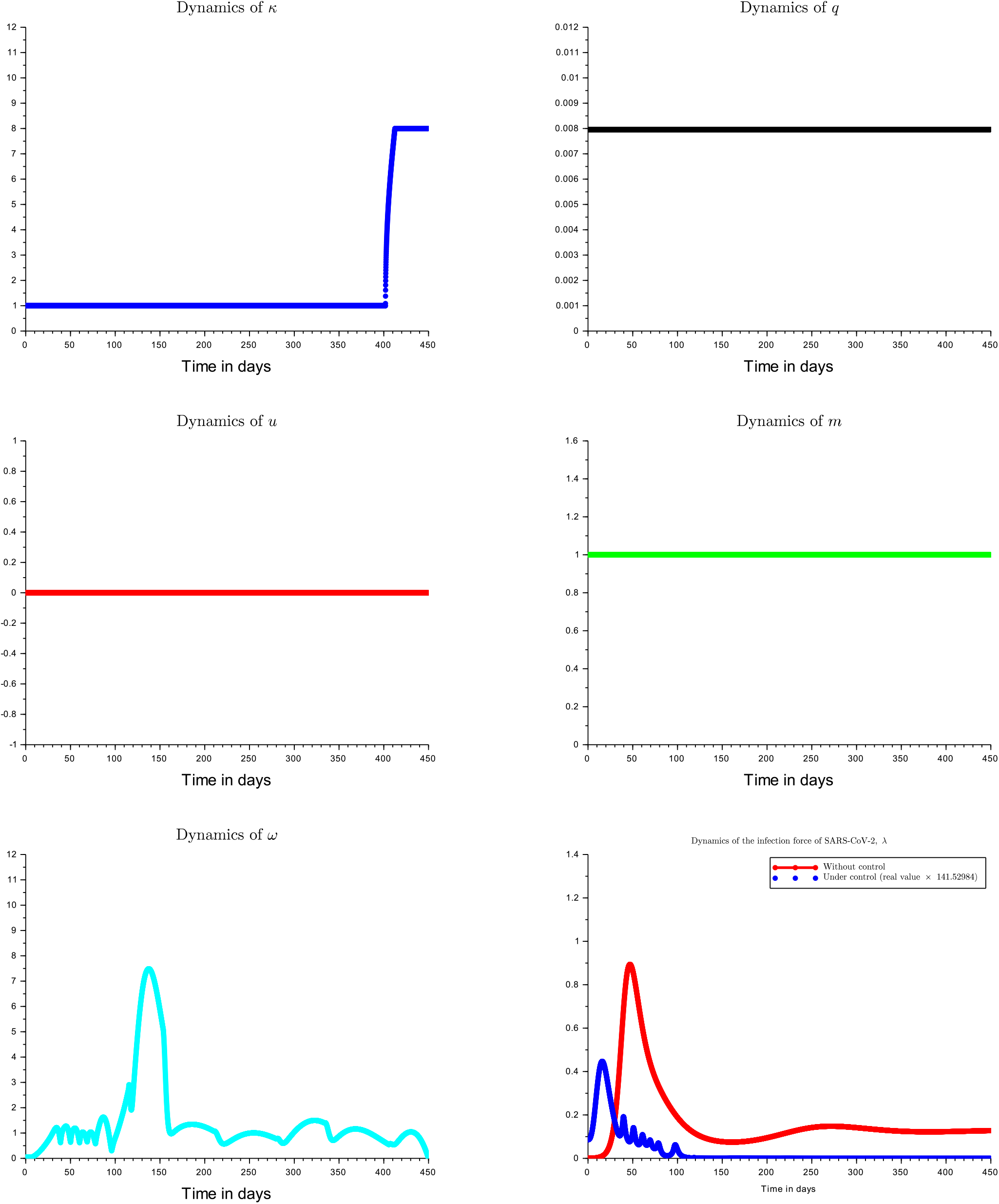
Optimal control dynamics of Covid-19 prevelence

Figure 6 shows the progression of the SARS-CoV-2 population (in terms of number of viruses per *m*^−2^) and the disease prevalence either the optimal control is applied or not. Relative prevalence of disease is preferred over absolute prevalence because it is more appropriate for scaling up. The peak of the disease occurs earlier and with smaller values under optimal control policy. We also observe that without control there is a possibility to observe several peaks of the disease. Without control, the peak of the disease (32% prevalence) occurs after 57 days (about 8.5 weeks) while under control it occurs after 26.37 days (about 4 weeks) with a weaker magnitude (0.4478% prevalence). With the initial compartmental population sizes given in Table 4 and under the optimal control strategy, the disease extinction in the human population is reached after 9.571*𝒜 ×*10^−3^ *day×m*^−2^ while it occurs in the virus population after 2.023*𝒜 ×*10^−1^ *day×m*^−2^. If we consider the maximal disease prevalence *𝒥* 1 and the virus population size (*N*_*V*_) is equal to its upper bound *C*_*V*_ given in Proposition 2.1, then the disease extinction in the human population is reached after 5.499*𝒜 ×* 10^3^ *day × m*^−2^. Similarly, the extinction in virus population is reached after 9.8321*𝒜 ×* 10^4^ *day × m*^−2^. Thus, the sooner the control measures are taken, the sooner the disease can disappear. The dependence of the disease extinction time with respect to the surface *𝒜* clearly highlights the importance of lock-down. As we can observe, the extinction can be very slower in the virus population than in the human population. The cost of the optimal strategy has been computed for 180 days corresponding to half of the year. It has been evaluated at *𝒜 Z*\* = 3.978 *USD×𝒜 ×m*^−2^ which corresponds on average to 0.0221 *USD×𝒜 ×m*^−2^*×day*^−1^.

**Table 4:** Estimation and baseline values of parameters of the model (21)

| Parameter | Value | Unit | Source |
| --- | --- | --- | --- |
| $a_1$ | $3.117558 \times 10^6$ | | Assumed |
| $a_2$ | $2.091775 \times 10^6$ | *virus* | [32] |
| $u_i$ | 0 | percentage | Assumed |
| $m_i$ | 1 | percentage | Assumed |
| $\kappa$ | $[5.003 \times 10^{-5}, 8]$ | $person \times m^{-2}$ | www.populationdata.net |
| $\varsigma_i^{min}$ | $3 \times 10^{-1}$ | percentage | Assumed |
| $\varsigma_i^{max}$ | 1 | percentage | Assumed |
| $\pi_i$ | $6.73 \times 10^{-1}$ | $virus \times person^{-1} day^{-1}$ | [32] |
| $\tau_{1,i}$ | $2.8 \times 10^{-2}$ | $day^{-1}$ | [32] |
| $\tau_{2,i}^{min}$ | 0 | $day^{-1}$ | Assumed |
| $\tau_{2,i}^{max}$ | $2.8 \times 10^{-2}$ | $day^{-1}$ | [32] |
| $\tau_{3,i}^{min}$ | 0 | $day^{-1}$ | Assumed |
| $\tau_{3,i}^{max}$ | $2.75 \times 10^{-1}$ | $day^{-1}$ | [32] |
| $\tau_4^{min}$ | 0 | $day^{-1}$ | Assumed |
| $\tau_4^{max}$ | 1 | $day^{-1}$ | [32] |
| $\nu$ | $\frac{1}{7}$ | $day^{-1}$ | [10] |
| $\omega_i$ | 0 | $day^{-1}$ | Assumed |
| $\mu$ | $\frac{1}{21681}$ | $day^{-1}$ | wikipedia.org, www.populationdata.net |
| $d$ | $3.714847 \times 10^{-3}$ | $day^{-1}$ | Fitted |
| $b_i$ | $3.193 \times 10^{-2}$ | $day^{-1}$ | www.populationdata.net |
| $\Lambda_i$ | 0 | $person \times day^{-1} m^{-2}$ | Assumed |
| $\alpha_1$ | $\frac{1}{14}$ | $day^{-1}$ | [39] |
| $\alpha_2$ | $4.913973 \times 10^{-5}$ | $day^{-1}$ | Assumed |
| $q$ | $7.960346 \times 10^{-3}$ | $day^{-1}$ | Fitted |
| $p$ | $9.776358 \times 10^{-1}$ | percentage | Fitted |
| $\gamma_1$ | $\frac{1}{7}$ | $day^{-1}$ | [39] |
| $\gamma_2$ | $\frac{1}{7}$ | $day^{-1}$ | [39] |
| $\gamma_3$ | $\frac{1}{7}$ | $day^{-1}$ | [39] |
| $\gamma_4$ | $\frac{1}{7}$ | $day^{-1}$ | [39] |
| $\ell$ | $2.74 \times 10^{-3}$ | $day^{-1}$ | [32] |
| $\rho_1$ | $\frac{1}{3}$ | $day^{-1}$ | Assumed |
| $\rho_2$ | 1 | $day^{-1}$ | Assumed |
| $\mathcal{R}_0$ | $1.0759773 \times 10^2$ | | Fitted |
| $\mathcal{T}_0$ | $4.4791419 \times 10^4$ | | Fitted |
| $S(0)$ | 7.987228 | $person \times m^{-2}$ | wikipedia.org, www.populationdata.net |
| $E(0)$ | $9.538403 \times 10^{-4}$ | $person \times m^{-2}$ | Fitted |
| $I(0)$ | $2.615628 \times 10^{-4}$ | $person \times m^{-2}$ | Fitted |
| $Q(0) = H(0)$ | 0 | $person \times m^{-2}$ | Assumed |
| $T(0) = R(0)$ | 0 | $person \times m^{-2}$ | Assumed |
| $D(0) = D_I(0) = B(0)$ | 0 | $person \times m^{-2}$ | Assumed |
| $Y_1(0)$ | $5.133665 \times 10^{-5}$ | $virus \times m^{-2}$ | Fitted |
| $Y_i(0) \ i = 2, \dots, 9$ | $5.133665 \times 10^{-5}$ | $virus \times m^{-2}$ | Assumed |
| $Y_{10}(0)$ | 0 | $virus \times m^{-2}$ | Assumed |

**Figure 6.**
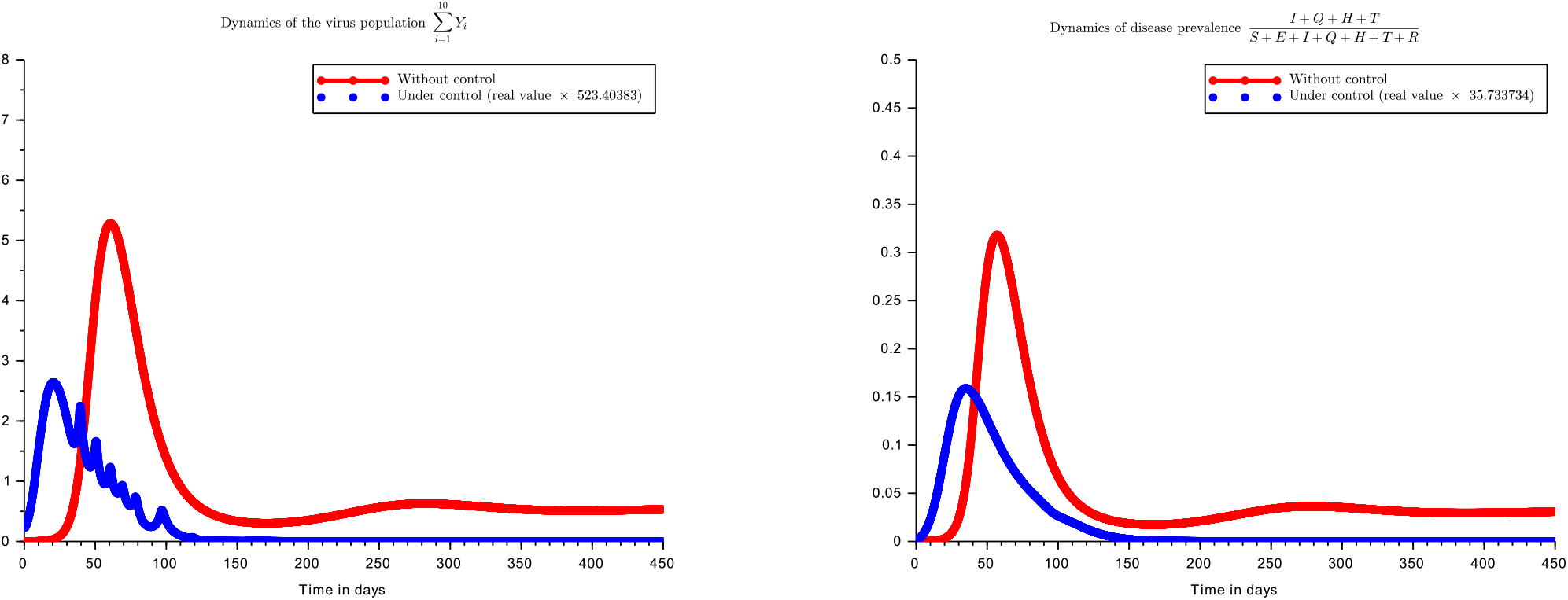
Controlled dynamics of Covid-19 prevalence

## 5 Conclusion

This work is concerned with the mathematical modeling of the severe acute respiratory corona virus 2. According to governmental data and World Health Organization, the corona virus made several millions of dead in the world. It spreads very rapidly and affects not only the public health sector, but also the global economy. The transmission of the disease can occur through direct contact with an infectious individual or through indirect contacts via the environment. The high complexity of the disease spreading suggested numerous modeling activities. Indeed, through a mathematical model it is possible to rigorously study fighting policies against corona. Hence, the goals we aimed to achieve were to propose a realistic model for the Covid-19 dynamics, to estimate the parameters of the proposed model according to the available real data, to study asymptotic behaviors depending on some featured control parameters related to WHO and governmental recommendations, and to design control strategies that are optimal with regards to socio-economical constraints.

We have constructed a two scale compartmental model with 20 classes corresponding to 10 human population states (macroscopic scale) and 10 viruses locations (microscopic scale). Indeed, we considered Susceptible (*S*), Infected (*E*), Infectious (*I*), Quarantined (*Q*), Hospitalized (*H*), Treated (*T*), Recovered (*R*), Non-Infectious dead (*D*), Infectious dead (*D*_*I*_), Buried (*B*). Each individual was assumed to have an external viral load on his body while there were free viruses in the environment. The model has been labeled 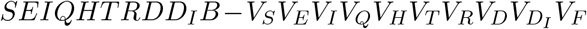 as described graphically in the Figure 1. To better access the effects of social distancing, the units of state variables have been taken in terms of person per unit area and virus per unit of surface. The parameters of the model have been successfully fitted using a Cameroonian dataset and a semi-empiric procedure. Additionally to the quantitative study of the system, we carried out a qualitative analysis of the model.

The existence of equilibriums has been studied. Contrarily to the endemic equilibrium, the disease free equilibrium (DFE) exists unconditionally. The DFE is globally asymptotically stable if a threshold *𝒯*_0_ is less or equal to one. We also provided a formula for the basic reproduction number *ℛ*_0_ which has a biological interpretation regarding the model: the average number of new viruses generated by an introduced virus during its lifespan. *𝒯*_0_ has a similar biological interpretation. A convergence index *𝒯*_1_ has been defined. Under an exponential convergence hypothesis, the times of disease extinction respectively in human population 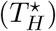 and virus population 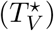 are determined. A sensitivity analysis of *ℛ*_0_, *𝒯*_0_ and *𝒯*_1_ have been done using the partial rank correlation coefficient (PRCC). We found that the sensitive parameters are in descending order the proportion of time of wearing mask, the proportion of time spent in disinfecting people and the environment, and the mobility probability. Through a concept of specific area *𝒜*, the importance of lock-down is highlighted. Indeed, the times of extinction are proportional to *𝒜* knowing that 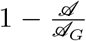 is the lock-down level. Since the disease and the application of the different control strategies generate economical costs, an optimal control analysis has been already carried out.

In order to guarantee effectiveness of control strategies and to reduce the expenses generated by Covid-19, we defined a functional cost to be minimized. The existence and the uniqueness of the optimal control is established and its characterization is given. By the sweep decent method the optimal strategies are computed according to some practical constraints materialized by an admissible control set. We found that if one globally considers the economical losses and gains, then no specific control effort is needed additionally to taking care of hospitalized patients and putting in quarantine of infectious reported individuals as it is already done. However, since the logistic resources for quarantine and hospitalization are limited, the economical losses in terms of gross domestic product (due to social distancing, reduction of mobility and lock-down) can be penalized in the functional cost. That has been done and we computed new optimal strategies.

According to the simulations, we observed that social distancing and disinfection frequency are the main elements of the optimal control strategy. Social distancing permits to have no restriction on mobility. The use of mask is useful but not necessary when social distancing and regular disinfection are respected. Screening has no significant effect on disease behavior and it is economically better to keep *q* at its minimal value corresponding to self decision to be tested or the hospitalization of severe disease cases. Under the optimal control policy, the peak of the disease occurs earlier and with smaller values.

The current work has several perspectives. The first one is to take into consideration the effects of a potential vaccine against Covid-19 both in the asymptotic behavior analysis and in the optimization of the control strategy. Another aspect is to consider the spatial heterogeneity in terms of available logistic resources, population densities, attractiveness of specific places at given periods of a day, cultural and religious habits, and others significant factors. An ongoing work is trying to tackle the problem regarding those aspects.

## Data Availability

Data are freely provided by the government

https://www.data.gouv.fr/fr/datasets/coronavirus-covid19-evolution-par-pays-et-dans-le-monde-maj-quotidienne/

https://www.humanitarianresponse.info/en/operations/cameroon/health/documents

## Acknowledgment

The authors would like to thank the anonymous editors and reviewers for their comments, which considerably improved the quality of the work.

## A Tables of state variables and parameters

### B Proofs of different results

**Proof of Proposition 2.1**.

Assume that the initial condition is taken in Ω. Using Cauchy-Lipschitz Theorem there exists a maximal solution of the system (21). Assume that the solution is defined and remains nonnegative on a set [0, *T*_*f*_]. Then, on]0, *T*_*f*_ [, *N* satisfies

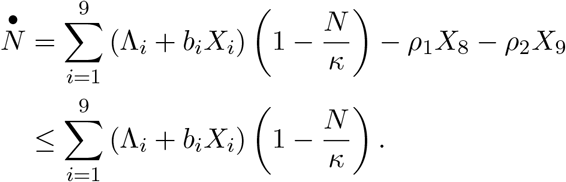

Thus, the orbit of *N* is clearly converges to the set]0, *κ*]. *X*_8_, *X*_9_ and *X*_10_ satisfy respectively

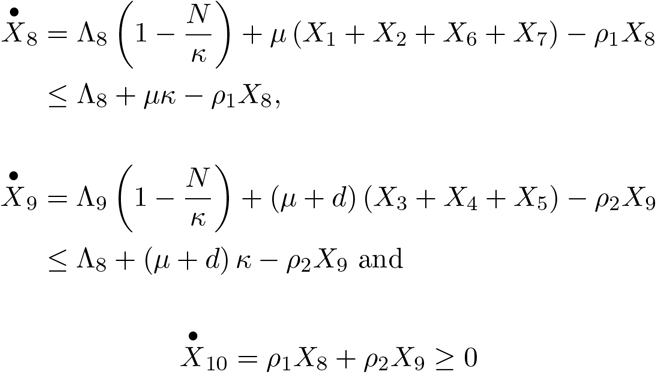

Hence, *X*_10_ ≥ 0 and the orbits of *X*_8_, *X*_9_ converge respectively to the sets [0, *C*_*D*_] and 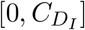. On the other hand,

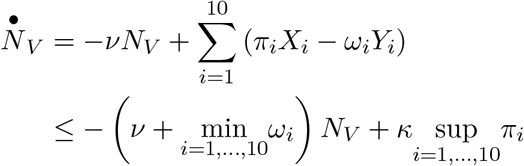

Looking carefully the system (1) − (20), we can see that 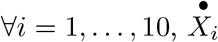 (respectively 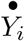) is nonnegative when *X*_*i*_ = 0 (respectively *Y*_*i*_ = 0). This proves the positivity of the solution which is finally global since it is bounded by the attractiveness of Ω.

**Proof of Proposition 2.2**.

i. Regarding the form of the model (21), it is plain that it admits the equilibrium given by *X* = *Y* = 0.
ii. Again, regarding the form of the model (21) the DFE exists if and only if *λ* = 0 and *b*_1_ (*κ* − *X*_1_) = *µκ*. Indeed, at the equilibrium 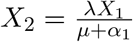. So it is necessary to have *λ* = 0.
iii. Once more, regarding the form of the model (21) the EE exists only if *λ* > 0 and

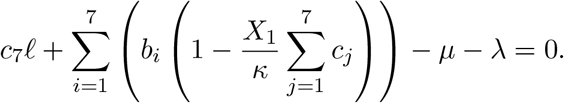

Solving the last equation leads to the expression of 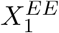 given above. The linear relation 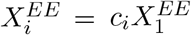 come from matrix *M*.

▪

**Proof of Proposition 2.3**.

i. Looking at the expression of *J* in {0}^9^ × ℝ_+_ × {0}^10^, we can see that *J*_1,2_ = 0 and both matrices *J*_1,1_ and *J*_2,2_ have all their eigenvalues negative except *J*_1,1_ (10, 10) which is null (the compartment of buried individuals is a sink). This shows that the set {0}^9^× ℝ_+_ × {0}^10^ is locally asymptotically stable (LAS) if *b*_1_ ≤ *µ*.
ii. If *b*_*i*_ ≤ *µ, i* = 1, *…*, 7 then it suffices to look at the equation of 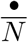 to get that the null equilibrium *X* = *Y* = 0 is GAS. Indeed,

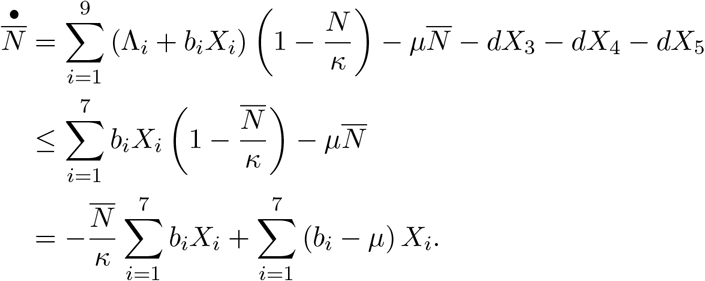 Since the *X*_*i*_s are nonnegative, if *b*_*i*_ ≤ *µ, i* = 1, *…*, 7 then 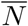 converges to 0 and necessarily *X* and *Y* converge to zero by construction of the model.
iii. From (*i*) other equilibrium than the null equilibrium is unstable if *b*_1_ ≤ *µ*. So, it is necessary to have *b*_1_ > *µ*. The disease free equilibrium is locally asymptotically stable (LAS) if *J* is Metzler stable. That is the case if *J*_1,1_, *J*_2,2_ and 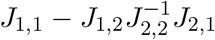 are all Metzler stable (see Proposition 3.1 in [31]). *J*_1,1_ and *J*_2,2_ are unconditionally Metzler stable. So it remains the stability of the matrix 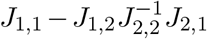.
iv. Assume that *b*_1_ > *µ* and *λ* = 0. Due to the linear form of equations (2) − (7), (9) we can easily see that *X*_*i*_, *i* = 2, *…*, 7, 9 tends to zero. Hence, regarding equations (1) and (8) we can check that *X*_1_ tends to 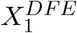 while *X*_8_ tends to 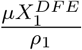.

▪

**Proof of Lemma 2.1**. The solution of (23) − (24) is given by

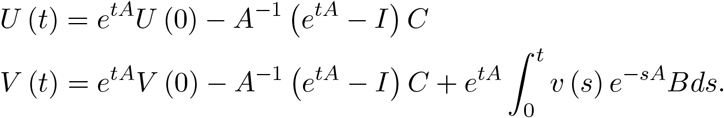

Let *α* (*A*) < 0 denote the stability modulus of the matrix *A*. We have

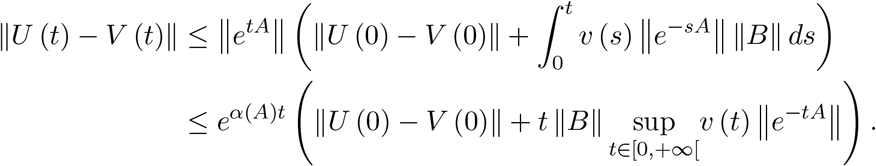

The results follows. ▪

**Proof of Theorem 2.1**. From Proposition 2.3, if *b*_1_ > *µ* and *λ* = 0 then the DFE is GAS. Using Lemma 2.1 the result is immediate. ▪

**Proof of Theorem 2.2**. The proof is based on the quasilinear form of the model (21). Indeed, the model (21) can be rewritten as

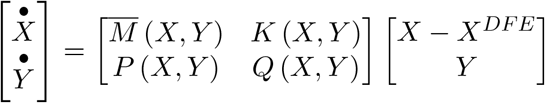

where 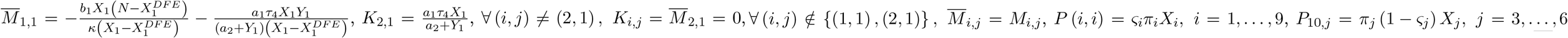. The model has an almost triangular form and is Metzler with a negative diagonal. The matrix 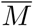 is Metzler stable by its particular form (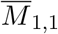 is a negative eigenvalue and the complementary diagonal block is triangular and Metzler stable). The matrix *Q* is Metzler and stable since its transpose is strictly diagonal dominant. Using Proposition 3.1 in [31], the stability is ensured if 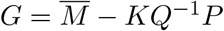 is Metzler stable. We have 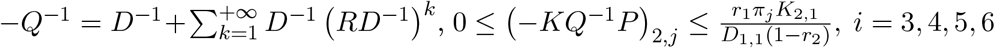 and (−*KQ*^−1^*P*)_*i,j*_*=* 0 otherwise. Hence, *G*_2,*j*_ = (−*KQ*^−1^*P*)_2,*j*_, *j* = 3, 4, 5, 6 and 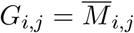 otherwise. *G* is therefore a Metzler. The stability of *G* is equivalent to the one of the sub-matrix (*G*_*i,j*_)_2≤ *i,j* ≤ 6_ due to the particular form of *G*. Again, using Proposition 3.1 in [31], the stability is ensured if

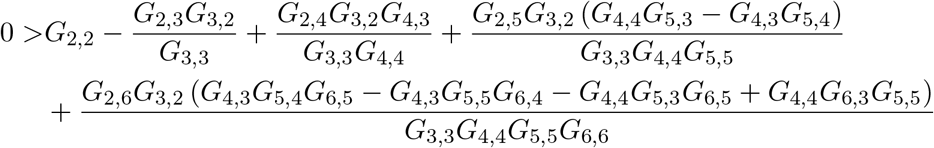

We have

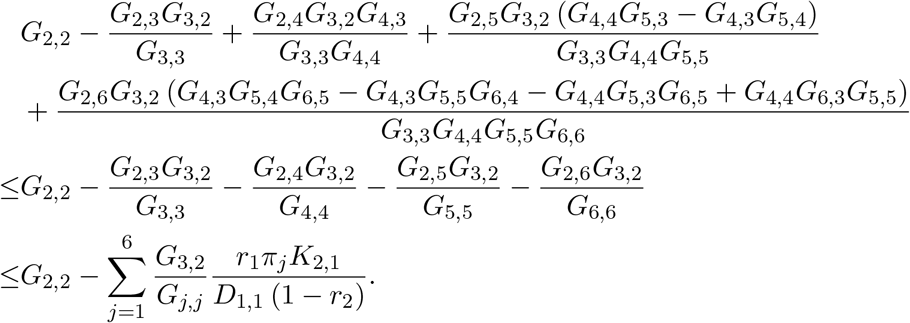

Thus, the stability holds if

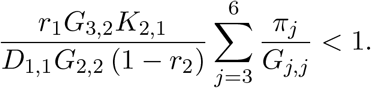

Using the boundedness of *X*_1_ and *N*_*V*_, we have 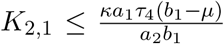 and we get the sufficient condition *𝒥*_0_ < 1. Since 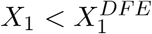 and 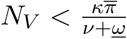, the stability holds if *𝒥*_0_ ≤ 1. ▪

**Proof of Theorem 2.3**. Let 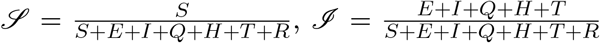 and assume that ∀*t* ≥ 0, *λ* (*t*) = *λ*_0_*e*^−*αt*^, *α* > 0. From the model (1)-(20) we have according to probabilistic properties of Poisson processes [8, 17, 33],

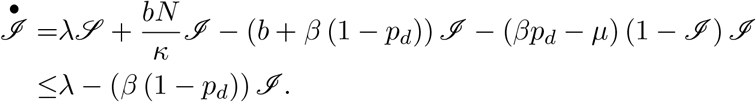

Indeed, the real phenomenon is a tuple of Poisson processes which jumps with rates given by different transition rates. The average time between two consecutive jumps is the inverse of the jumping rate and almost surely only one process in the tuple jumps at a given time. Hence, the average time 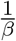 spent by an individual from his entry in the compartment *E* to his exit from the compartment *T* is the sum of average times he spent in of each the compartment *E, I, Q, H* and *T* respectively. If *σ*_1_ = min {*α, β* (1 − *p*_*d*_)} then using the resolvent operator we have ∀*t* ≥ 0,

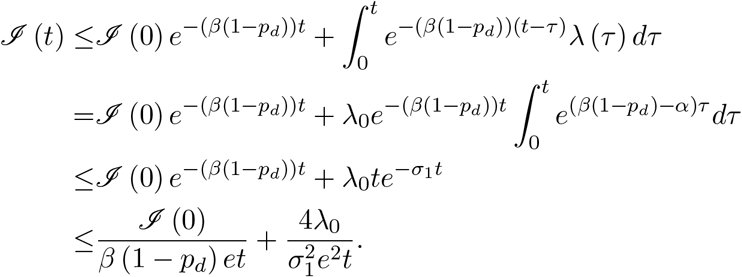

The last inequality comes from the fact that 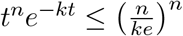 for *k, n* ∈ ℕ^*^ and *t* ∈ ℝ_+_. Since 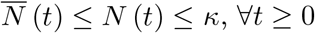, we have

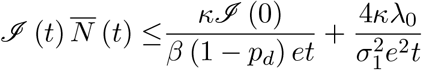

Let σ_2_ = min {*v* + *<u>w</u>, β* (1–*p*_*d*_)} and σ_3_ = min {*v* + *<u>w</u>*, σ_1_}. We have

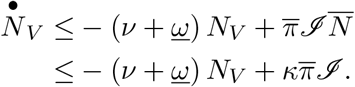

Again, using the Gronwall’s Lemma we have

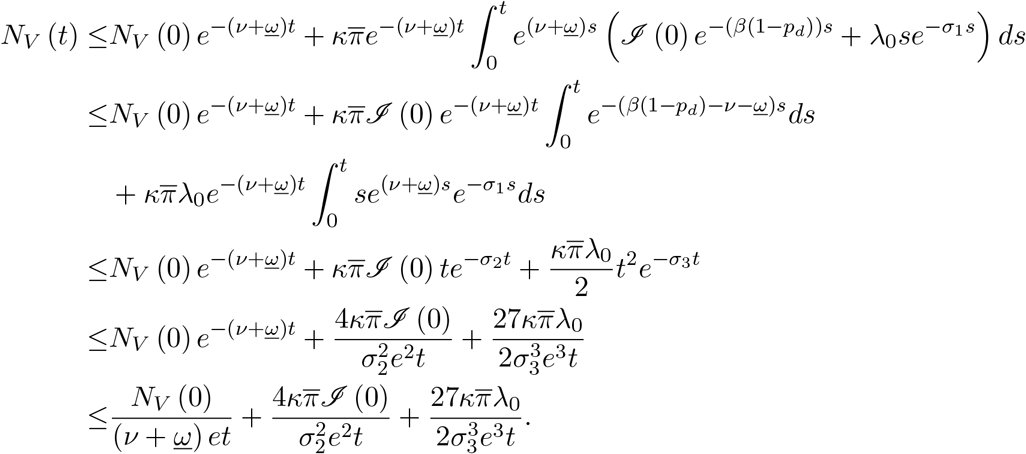

Hence, the respective first times 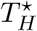 and 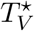 such that respectively 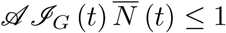 and *𝒜 N*_*V*_ (*t*) ≤ 1 satisfy

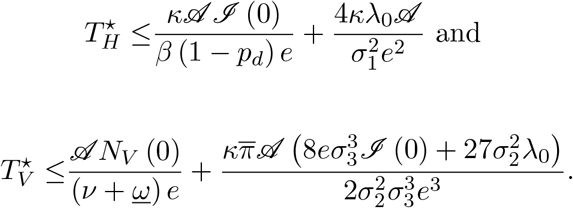

▪

**Proof of Theorem 4.1**. The existence of the optimal control is guaranteed by the boundedness of the solution of (1)-(20) and subsequently, the boundedness of the functional cost *Z* which is continuously differentiable with respect to its parameters. The rest of the proof is essentially based on the study of the gradient of the Hamiltonian. Indeed, the minimization consists in moving according to the opposite direction of the gradient of the Hamiltonian while it is possible and necessary. Precisely, when the gradient is the null vector then a singular point is reached. If the gradient is not zero but it is not possible to move again, then the optimum is reached at the boundary of the eligible region ^7^. We have

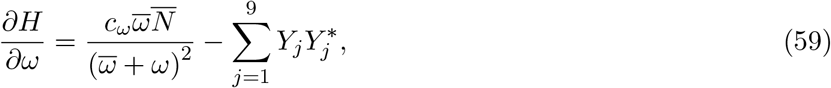

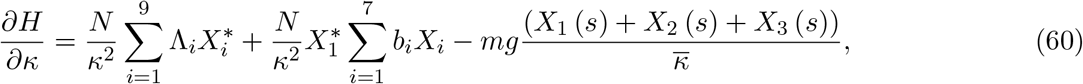

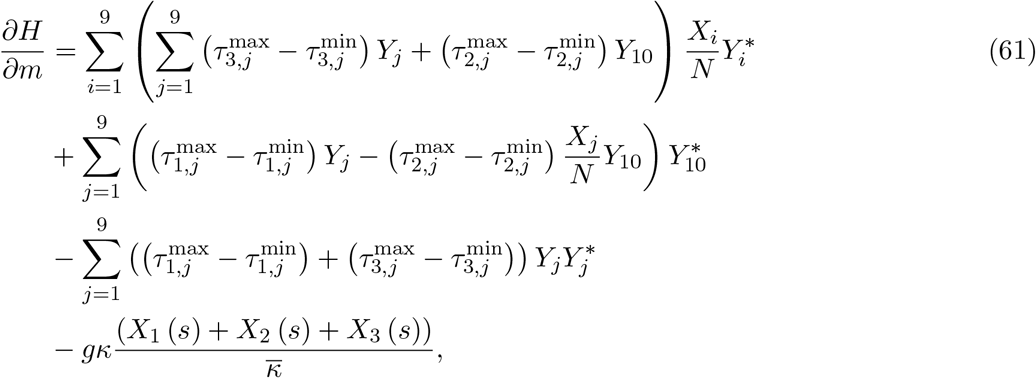

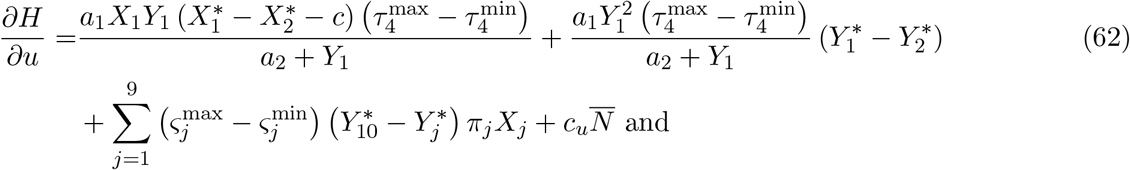

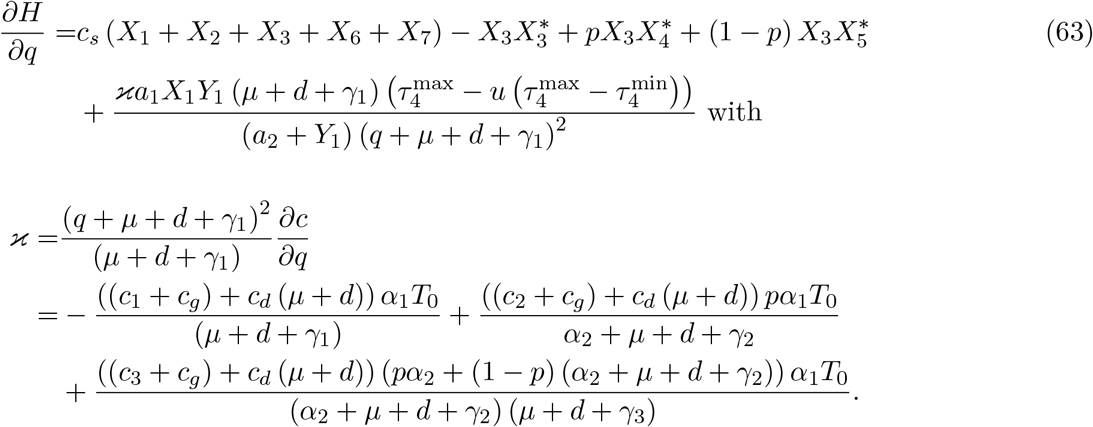

If the optimal control exists then the Algorithm 1 describes it such as a piecewise continuous feedback which is locally Lipschitz with respect to the state variable and the adjoint state variable. Thus, by the Cauchy-Lipschitz Theorem the optimal control is unique.▪

## C Computation of the optimal control

### Algoritham 1: Optimal control (part 1)

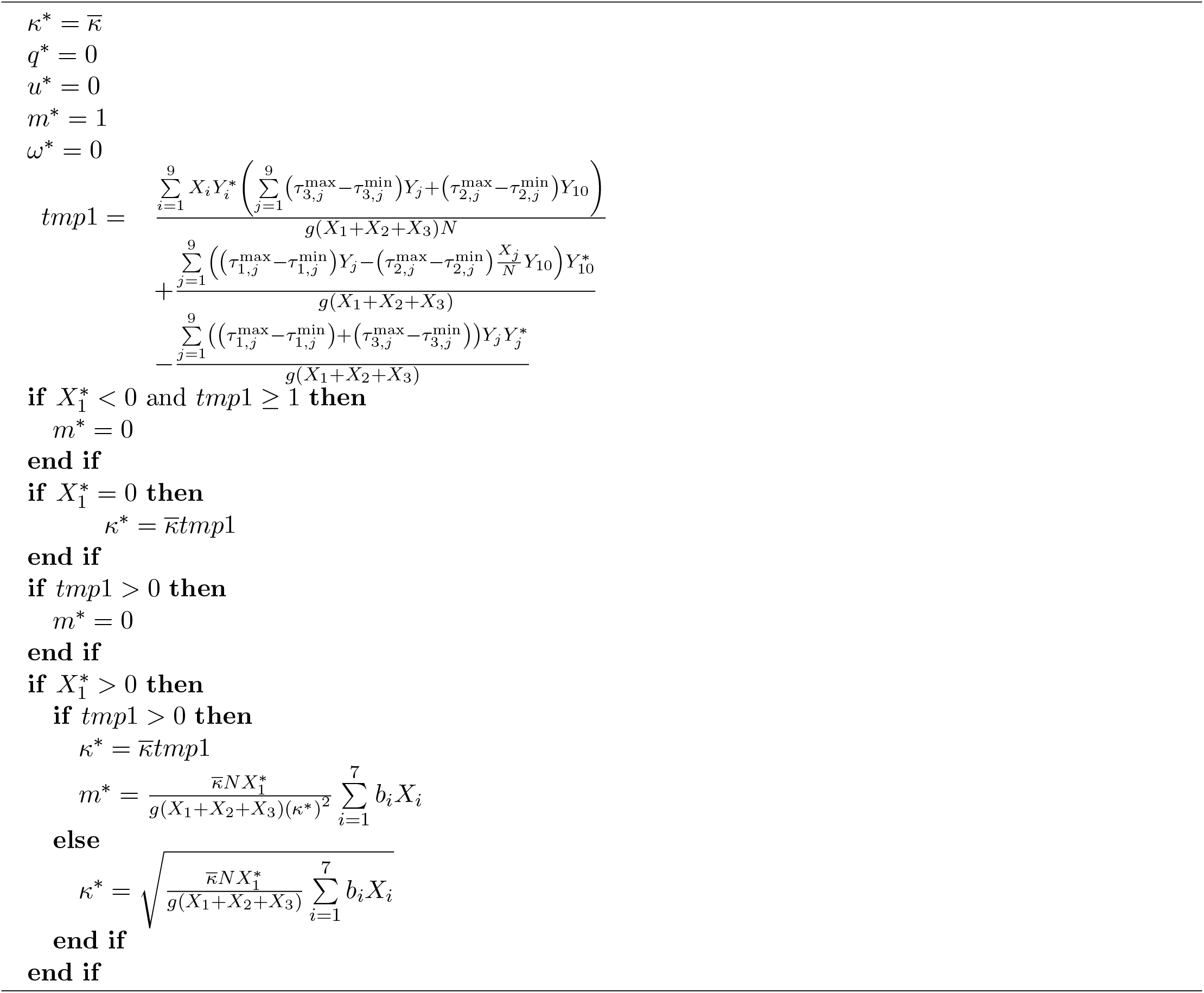

### Algoritham 2: Optimal control (part 2)

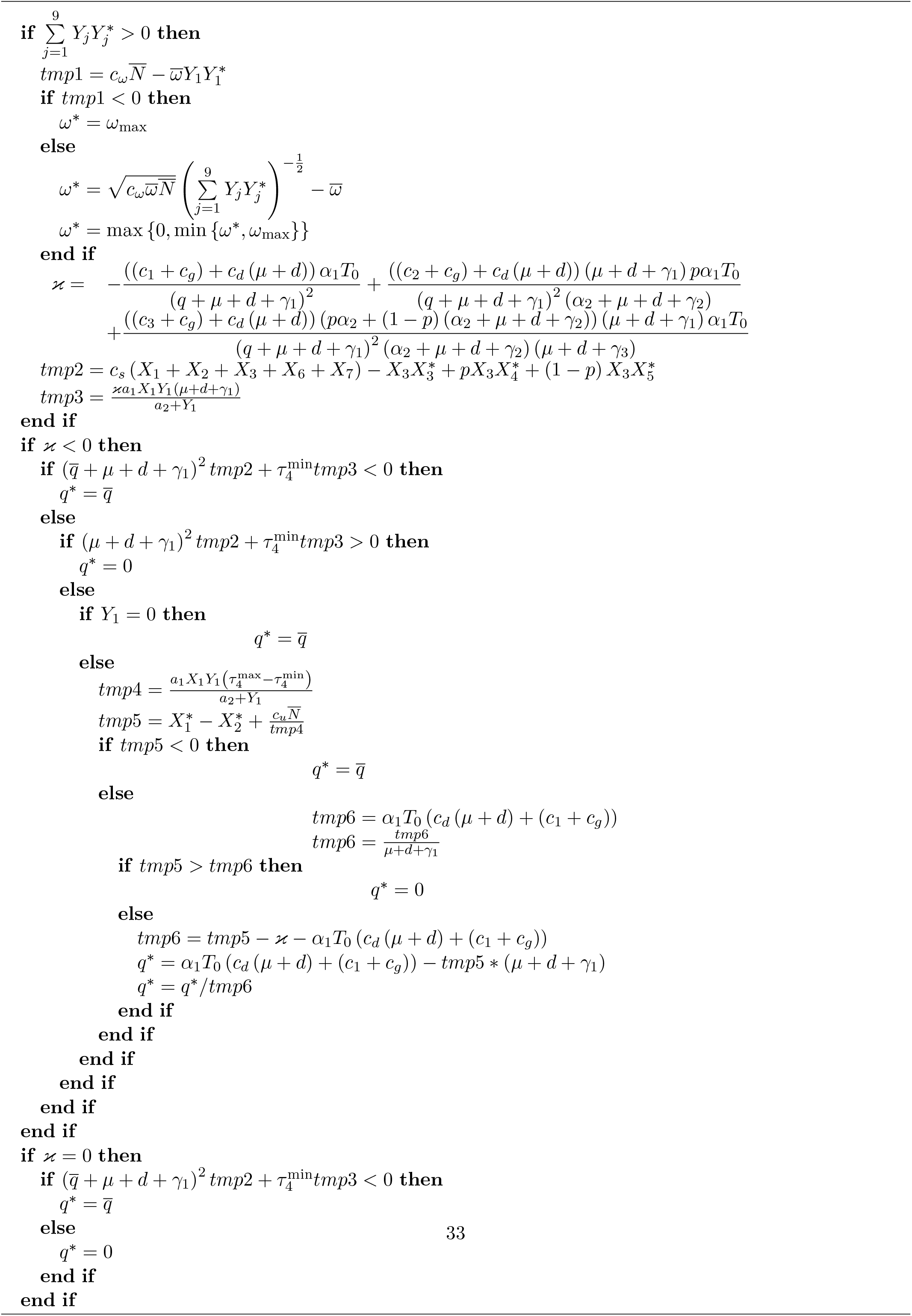

### Algoritham 3: Optimal control (part 3)

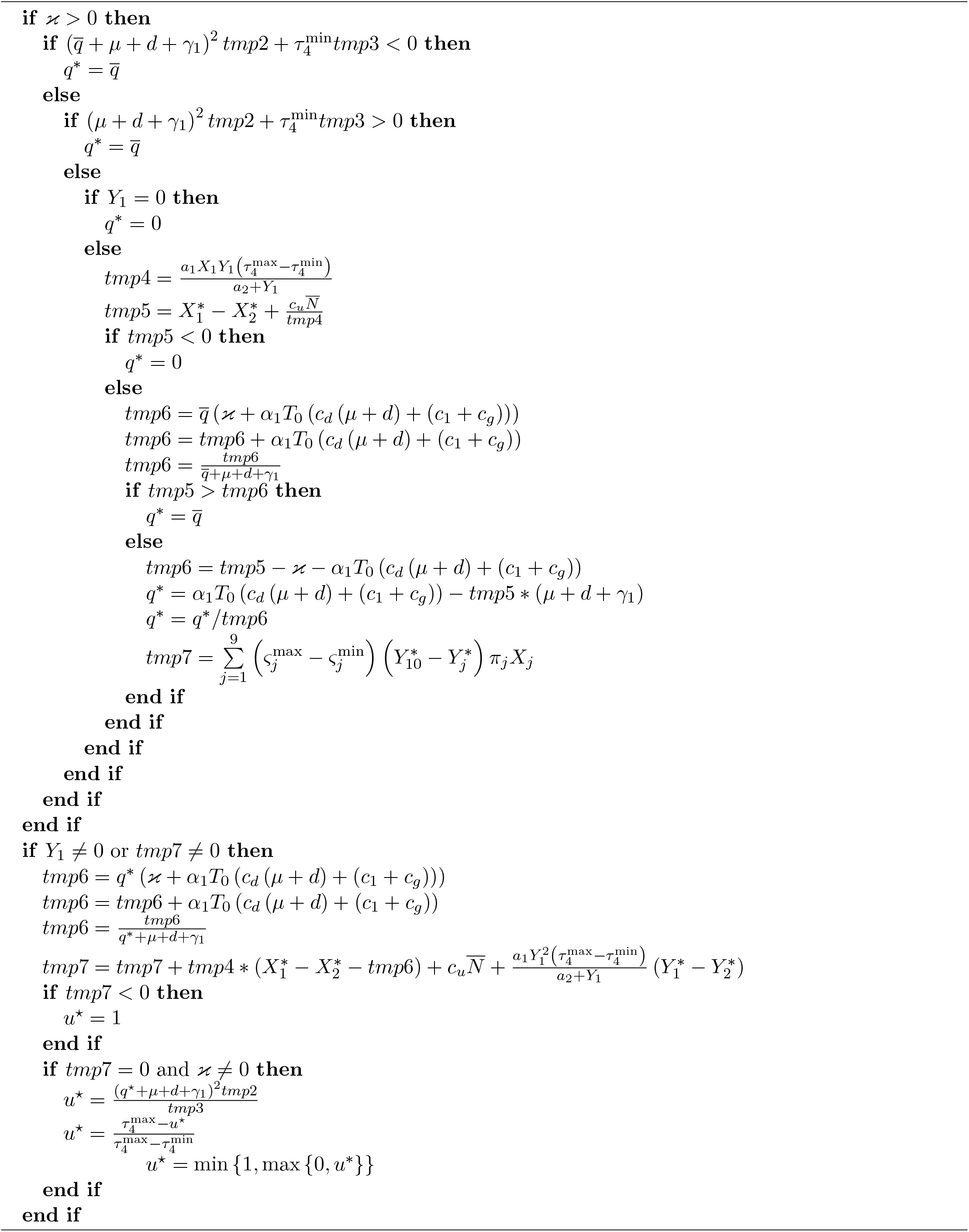

## Footnotes

1 https://www.who.int/emergencies/diseases/novel-coronavirus-2019

2 See https://medicalaid.org

3 https://www.cdc.gov/coronavirus/

4 See the book [27] for more details.

5 www.populationdata.net

6 www.populationdata.net

7 See the book [2] for more details.

## References

[1] T. Alberti and D. Faranda. On the uncertainty of real-time predictions of epidemic growths: a covid-19 case study for china and italy. Communications in Nonlinear Science and Numerical Simulation, page 105372, 2020.

[2] S. Anita, V. Capasso, and V. Arnautu. An Introduction to Optimal Control Problems in Life Sciences and Economics: From Mathematical Models to Numerical Simulation with MATLAB 2011. OR. Springer,

[3] M. Batista. Estimation of the final size of the second phase of Coronavirus epidemic by the logistic model. MedRxiv, 2020.

[4] S. Bekiros and D. Kouloumpou. SBDiEM: A new mathematical model of infectious disease dynamics. Chaos, Solitons & Fractals, page 109828, 2020.

[5] Y. Belgaid, M. Helal, and E. Venturino. Analysis of a model for Coronavirus spread. Mathematics, 8(5):820, 2020.

[6] S. Bowong and J. Kurths. Modeling and parameter estimation of Tuberculosis with application to cameroon. International Journal of Bifurcation and Chaos, 21(07):1999–2015, 2011.

[7] S. Bowong, L. Mountaga, A. Bah, J. Tewa, and J. Kurths. Parameter and state estimation in a Neisseria meningitidis model: A study case of niger. Chaos: An Interdisciplinary Journal of Nonlinear Science, 26(12):123115, 2016.

[8] T. Britton, E. Pardoux, F. Ball, C. Laredo, D. Sirl, and V. C. Tran. Stochastic epidemic models with inference. Springer, 2019.

[9] C. Castilho, J. A. Gondim, M. Marchesin, and M. Sabeti. Assessing the efficiency of different control strategies for the covid-19 epidemic. Electronic Journal of Differential Equations, 64(2020):1–17, 2020.

[10] A. Chin, J. Chu, M. Perera, K. Hui, H.-L. Yen, M. Chan, M. Peiris, and L. Poon. Stability of SARS-CoV-2 in different environmental conditions. MedRxiv, 2020.

[11] N. Chitnis, J. M. Hyman, and J. M. Cushing. Determining important parameters in the spread of malaria through the sensitivity analysis of a mathematical model. Bulletin of mathematical biology, 70(5):1272, 2008.

[12] R. Djidjou-Demasse, Y. Michalakis, M. Choisy, M. T. Sofonea, and S. Alizon. Optimal COVID-19 epidemic control until vaccine deployment. MedRxiv, 2020.

[13] S. E. Eikenberry, M. Mancuso, E. Iboi, T. Phan, K. Eikenberry, Y. Kuang, E. Kostelich, and A. B. Gumel. To mask or not to mask: Modeling the potential for face mask use by the general public to curtail the COVID-19 pandemic. Infectious Disease Modelling, 2020.

[14] B. Espinoza, C. Castillo-Chavez, and C. Perrings. Mobility restrictions for the control of epidemics: When do they work? PloS one, 15(7):e0235731, 2020.

[15] D. Fanelli and F. Piazza. Analysis and forecast of COVID-19 spreading in China, Italy and France. Chaos, Solitons & Fractals, 134:109761, 2020.

[16] D. Faranda, I. P. Castillo, O. Hulme, A. Jezequel, J. S. Lamb, Y. Sato, and E. L. Thompson. Asymptotic estimates of SARS-CoV-2 infection counts and their sensitivity to stochastic perturbation. Chaos: An Interdisciplinary Journal of Nonlinear Science, 30(5):051107, 2020.

[17] D. Foata and A. Fuchs. Processus stochastiques: Processus de Poisson, chaînes de Markov et martin-gales. 2002.

[18] A. Fortin. Analyse numérique pour ingénieurs. Presses inter Polytechnique, 2011.

[19] R. B. Garabed, A. Jolles, W. Garira, C. Lanzas, J. Gutierrez, and G. Rempala. Multi-scale dynamics of infectious diseases, 2020.

[20] W. Garira. A complete categorization of multiscale models of infectious disease systems. Journal of biological dynamics, 11(1):378–435, 2017.

[21] W. Garira. The research and development process for multiscale models of infectious disease systems. PLoS computational biology, 16(4):e1007734, 2020.

[22] W. Garira and M. C. Mafunda. From individual health to community health: towards multiscale modeling of directly transmitted infectious disease systems. Journal of Biological Systems, 27(01):131– 166, 2019.

[23] W. Garira, D. Mathebula, and R. Netshikweta. A mathematical modelling framework for linked within-host and between-host dynamics for infections with free-living pathogens in the environment. Mathematical biosciences, 256:58–78, 2014.

[24] M. Gilbert, G. Pullano, F. Pinotti, E. Valdano, C. Poletto, P.-Y. Böelle, E. d’Ortenzio, Y. Yazdanpanah, S. P. Eholie, M. Altmann, et al. Preparedness and vulnerability of African countries against importations of COVID-19: a modelling study. The Lancet, 395(10227):871–877, 2020.

[25] T. Götz and P. Heidrich. Early stage COVID-19 disease dynamics in Germany: models and parameter identification. Journal of Mathematics in Industry, 10(1):1–13, 2020.

[26] J. Goupy and L. Creighton. Introduction aux plans d’expériences, volume 3. Dunod Paris, 2006.

[27] J. Harmand, C. Lobry, A. Rapaport, and T. Sari. The Chemostat: Mathematical Theory of Microorganism Cultures. John Wiley & Sons, 2017.

[28] J. Hilton and M. J. Keeling. Estimation of country-level basic reproductive ratios for novel Coronavirus (SARS-CoV-2/COVID-19) using synthetic contact matrices. PLOS Computational Biology, 16(7):e1008031, 2020.

[29] Z. Hu, Q. Cui, J. Han, X. Wang, E. Wei, and Z. Teng. Evaluation and prediction of the COVID-19 variations at different input population and quarantine strategies, a case study in Guangdong province, China. International Journal of Infectious Diseases, 2020.

[30] F. Jedrzejewski. Introduction aux méthodes numériques. Springer Science & Business Media, 2005.

[31] J. C. Kamgang and G. Sallet. Computation of threshold conditions for epidemiological models and global stability of the disease-free equilibrium (DFE). Mathematical biosciences, 213(1):1–12, 2008.

[32] S. M. Kassa, J. B. Njagarah, and Y. A. Terefe. Analysis of the mitigation strategies for COVID-19: from mathematical modelling perspective. Chaos, Solitons & Fractals, page 109968, 2020.

[33] J. Kingman. Poisson Processes. Oxford University Press, 1993.

[34] A. Kouidere, B. Khajji, A. El Bhih, O. Balatif, and M. Rachik. A mathematical modeling with optimal control strategy of transmission of COVID-19 pandemic virus. Commun. Math. Biol. Neurosci., 2020:Article–ID, 2020.

[35] D. La Torre, T. Malik, and S. Marsiglio. Optimal control of prevention and treatment in a basic macroeconomic-epidemiological model. Mathematical Social Sciences, 2020.

[36] S. Lenhart and J. T. Workman. Optimal control applied to biological models. CRC press, 2007.

[37] M.-T. Li, G.-Q. Sun, J. Zhang, Y. Zhao, X. Pei, L. Li, Y. Wang, W.-Y. Zhang, Z.-K. Zhang, and Z. Jin. Analysis of COVID-19 transmission in Shanxi province with discrete time imported cases. Mathematical Biosciences and Engineering, 17(4):3710, 2020.

[38] Z. Liu, P. Magal, O. Seydi, and G. Webb. A COVID-19 epidemic model with latency period. Infectious Disease Modelling, 2020.

[39] Z. Liu, P. Magal, O. Seydi, and G. Webb. Understanding unreported cases in the COVID-19 epidemic outbreak in Wuhan, China, and the importance of major public health interventions. Biology, 9(3):50, 2020.

[40] P. Magal and G. Webb. The parameter identification problem for SIR epidemic models: identifying unreported cases. Journal of mathematical biology, 77(6-7):1629–1648, 2018.

[41] M. Mandal, S. Jana, S. K. Nandi, A. Khatua, S. Adak, and T. Kar. A model based study on the dynamics of COVID-19: Prediction and control. Chaos, Solitons & Fractals, page 109889, 2020.

[42] S. Marino, I. B. Hogue, C. J. Ray, and D. E. Kirschner. A methodology for performing global uncertainty and sensitivity analysis in systems biology. Journal of theoretical biology, 254(1):178–196, 2008.

[43] M. Martcheva, N. Tuncer, and C. St Mary. Coupling within-host and between-host infectious diseases models. Biomath, 4(2):1510091, 2015.

[44] K. Nah, S. Chen, Y. Xiao, B. Tang, N. Bragazzi, J. Heffernan, A. Asgary, N. Ogden, and J. Wu. Scenario tree and adaptive decision making on optimal type and timing for intervention and social-economic activity changes to manage the covid-19 pandemic. European Journal of Pure and Applied Mathematics, 13(3):710–729, 2020.

[45] C. N. Ngonghala, E. Iboi, S. Eikenberry, M. Scotch, C. R. MacIntyre, M. H. Bonds, and A. B. Gumel. Mathematical assessment of the impact of non-pharmaceutical interventions on curtailing the 2019 novel Coronavirus. Mathematical Biosciences, page 108364, 2020.

[46] C. N. Ngonghala, E. A. Iboi, and A. B. Gumel. Could masks curtail the post-lockdown resurgence of COVID-19 in the US? MedRxiv, 2020.

[47] C. H. Nkwayep, S. Bowong, J. Tewa, and J. Kurths. Short-term forecasts of the COVID-19 pandemic: study case of cameroon. Chaos, Solitons & Fractals, page 110106, 2020.

[48] K. Prem, A. R. Cook, and M. Jit. Projecting social contact matrices in 152 countries using contact surveys and demographic data. PLoS computational biology, 13(9):e1005697, 2017.

[49] F. A. Rabi, M. S. Al Zoubi, G. A. Kasasbeh, D. M. Salameh, and A. D. Al-Nasser. SARS-CoV-2 and Coronavirus disease 2019: what we know so far. Pathogens, 9(3):231, 2020.

[50] Q. Richard, S. Alizon, M. Choisy, M. T. Sofonea, and R. Djidjou-Demasse. Age-structured non-pharmaceutical interventions for optimal control of COVID-19 epidemic. MedRxiv, 2020.

[51] W. C. Roda, M. B. Varughese, D. Han, and M. Y. Li. Why is it difficult to accurately predict the COVID-19 epidemic? Infectious Disease Modelling, 2020.

[52] K. J. Rothman, S. Greenland, and T. L. Lash. Modern epidemiology. Lippincott Williams & Wilkins, 2008.

[53] T. Sardar, S. S. Nadim, S. Rana, and J. Chattopadhyay. Assessment of lockdown effect in some states and overall India: A predictive mathematical study on COVID-19 outbreak. Chaos, Solitons & Fractals, page 110078, 2020.

[54] N. R. Sasmita, M. Ikhwan, S. Suyanto, and V. Chongsuvivatwong. Optimal control on a mathematical model to pattern the progression of coronavirus disease 2019 (covid-19) in indonesia. Global Health Research and Policy, 5(1):1–12, 2020.

[55] M. Serhani and H. Labbardi. Mathematical modeling of COVID-19 spreading with asymptomatic infected and interacting peoples. 2020.

[56] P. van den Driessche. Reproduction numbers of infectious disease models. Infectious Disease Modelling, 2(3):288–303, 2017.

[57] P. Van den Driessche and J. Watmough. Reproduction numbers and sub-threshold endemic equilibria for compartmental models of disease transmission. Mathematical biosciences, 180(1-2):29–48, 2002.

[58] P. Van den Driessche and J. Watmough. Further notes on the basic reproduction number. In Mathematical Epidemiology, pages 159–178. Springer, 2008.

[59] V. Volpert, M. Banerjee, A. dOnofrio, T. Lipniacki, S. Petrovskii, and V. C. Tran. Coronavirus-Scientific insights and societal aspects, 2020.

[60] Q. Wang, S. Xie, Y. Wang, and D. Zeng. Survival-convolution models for predicting COVID-19 cases and assessing effects of mitigation strategies. MedRxiv, 2020.

[61] X. Zhang, R. Ma, and L. Wang. Predicting turning point, duration and attack rate of COVID-19 outbreaks in major Western countries. Chaos, Solitons & Fractals, page 109829, 2020.

[62] L. Zhou, K. Wu, H. Liu, Y. Gao, and X. Gao. CIRD-F: Spread and influence of COVID-19 in china. Journal of Shanghai Jiaotong University (Science), 25:147–156, 2020.

